# Humoral immunity to endemic coronaviruses in older adults increases post-COVID-19 vaccination and in correlation with their anti-SARS-CoV-2 responses

**DOI:** 10.1101/2025.08.26.25334476

**Authors:** Sarah E. Woodward, Brynn McMillan, Gabrielle N. Gaultier, Ana Citlali Márquez, Tahereh Valadbeigy, Karen Simmons, Aidan M. Nikiforuk, Sofia R. Bartlett, Inna Sekirov, Muhammad Morshed, Manish Sadarangani, Agatha N. Jassem

## Abstract

Advanced age is an established risk factor for SARS-CoV-2 infection and severe disease. Older adults, having experienced recurrent infections with human endemic coronaviruses (HCoVs) throughout life, may have developed humoral immune responses that impact IgG targeting of both severe acute respiratory syndrome coronavirus (SARS-CoV) −2 and HCoVs. We profiled IgG responses in community-dwelling adults aged 50-87 years from the “PRospEctiVe EvaluatioN of immunity after COVID-19 vaccines” (PREVENT-COVID) prospective cohort study. Dried blood spot sampling occurred at baseline, prior to detected SARS-CoV-2 antigen exposure, and throughout the recommended vaccination schedule in Canada for SARS-CoV-2 seronegative and seropositive individuals. We define an HCoV-OC43-IgG high and HCoV-NL63-low baseline antibody landscape, which was unaffected by age or sex. Following COVID-19 vaccination, we observed increased anti HCoV-OC43 and HCoV-HKU1 IgG antibody titers, with substantial waning between doses. IgG specific to beta-coronaviruses and SARS-CoV-2 spike antigens positively correlated, strengthening with subsequent doses. In contrast, SARS-CoV-2 anti-nucleocapsid (N) antibodies associated most closely with HCoV-NL63 IgG levels. These findings suggest that HCoV-specific humoral immunity induced by COVID-19 vaccination or disease may impact susceptibility to future coronavirus infection. ACE2-binding assays revealed that individuals with greater HCoV-OC43- and HCoV-HKU1-specific IgG levels had the highest percent neutralization of the wildtype virus and SARS-CoV-2 variants. Together, our study highlights potential effects of COVID-19 vaccination on humoral immunity towards related coronaviruses.

## Introduction

Coronaviruses (CoV) have long been understood to harbor pandemic potential, as evidenced by previous outbreaks of severe acute respiratory syndrome coronavirus (SARS-CoV) −1 and Middle East respiratory syndrome (MERS) coronavirus (1). However, of the seven coronaviruses currently known to infect and cause disease in humans, four are now considered endemic: 229E, NL63, OC43, and HKU1. Of these, 229E and NL63 belong to the family alphacoronaviridae, and OC43 and HKU1 belong to the family betacoronaviridae and are more closely related to SARS-CoV-2, the causative agent of coronavirus disease-2019 (COVID-19) (1).

Prior to the COVID-19 pandemic beginning in late 2019, endemic coronaviruses were responsible for between 5-30% of upper respiratory infections such that 90-100% of individuals are estimated to have developed some degree of humoral immunity against human endemic coronaviruses (HCoVs) by ten years of age (2). Despite this widespread community-level immunity towards HCoVs, reinfection occurs approximately every two to three years in the adult population (3), suggesting that immune waning occurs. The COVID-19 pandemic began in the context of widespread circulation of endemic coronaviruses, and consequently, of variable population-wide immunity to viruses closely related to SARS-CoV-2.

Studies have now identified a general relationship between SARS-CoV-2 antigen exposure, via natural infection or vaccination, and increased levels of antibodies targeting HCoVs, as well as cross-reactivity of human anti-coronavirus antibodies to multiple related viral lineages (4–6). However, mathematical modelling has also suggested that life-long, repeated, exposure to HCoVs could blunt immunity to SARS-CoV-2 by promoting expansion of antibodies specific to previously exposed coronavirus strains (so-called “back-boosting”), rather than antibodies against SARS-CoV-2, in a phenomenon known as “immune imprinting” (7). Furthermore, high antibody concentrations do not always correlate with increased protection from viral infection. Therefore, questions remain regarding the relationship between prior endemic coronavirus exposure, the humoral response to COVID-19 vaccination, and overall susceptibility to SARS-CoV-2 infection in the older adult population. Particularly, whether anti-HCoV antibody titres influence vaccine or infection-induced anti-SARS-CoV-2 immunity.

To understand the relationship between HCoV and SARS-CoV-2 humoral immunity, we performed a prospective cohort study in community-dwelling adults aged 50 and older in British Columbia, Canada. Antibody responses targeting HCoVs 229E, NL63, OC43 and HKU1, as well as SARS-CoV-1 and SARS-CoV-2, were profiled over the course of the recommended COVID-19 vaccination schedule. We characterized baseline anti-HCoV antibody levels, prior to known exposure to SARS-CoV-2 antigens, and demonstrated that COVID-19 vaccination boosts HCoV-specific IgG levels in both SARS-CoV-2 seronegative and seropositive individuals. A positive correlation was identified between anti-OC43 and anti-HKU1 IgG levels with both SARS-CoV-2 anti-Spike (S) antibody titres and viral neutralization activity. Overall, this work provides insights into coronavirus-specific IgG antibody dynamics post-vaccination in older adults, towards better understanding the relationship between HCoV and SARS-CoV-2 humoral immunity.

## Methods

### Ethics

Ethics approval for the “PRospEctiVe EvaluatioN of immunity after COVID-19 vaccines” (PREVENT-COVID) study was provided by the University of British Columbia Clinical Research Ethics Board (Ref: H20-03951). Written informed consent was obtained from all study participants. Further data was used from the Accelerating SARS-CoV-2 Seroprevalence SurveyS through Dried Blood Spots (ASSESS-DBS) study (8). Ethics approval for the ASSESS-DBS study was granted by the University of British Columbia Clinical Research Ethics Board (Ref: H20-02402).

### Cohort recruitment and participant demographics

Participants were enrolled in the study PREVENT-COVID, full details of which are included in McMillan B *et al* (9). Adults aged ≥19 years who were planning to receive a COVID-19 vaccine were eligible for the study. Participants were excluded if: presence of a bleeding disorder; condition resulting in a compromised immune system; taking medication during the study time course that could affect immune responses to vaccination. The present analysis was restricted to those aged ≥50 years at the time of the first study visit. Vaccines were provided outside the study, by the regular immunization provider. A total of 611 individuals over the age of 50 were included in this analysis, after further filtering of PREVENT-COVID study participants who had complete demographic information.

Demographic data including age, sex, and self-reported ethnicity were recorded and participants self-reported the presence or absence of comorbidities as well as prior SARS-CoV-2 infection status. Data used in this study were obtained from survey responses and analysis of reconstituted dried blood spot (DBS) samples self-collected via fingerprick over the course of nine visits spanning from February 2021 to August 2023, with study visits chosen to reflect key time points post-COVID-19 vaccination. To capture asymptomatic SARS-CoV-2 infection status, patient serology was further tested for the presence of anti-nucleocapsid (N) antibody, indicative of infection-induced immunity in combination with a positive anti-spike response, as the N protein was not included in Health Canada-approved vaccines (see ‘Serological testing and analysis of dried blood spot samples’). Symptomatic SARS-CoV-2 infection status was further reported via linkage to the provincial testing database for indication of positive COVID-19 PCR test results (10).

### Sample collection and DBS preservation

Samples were collected as described by McMillan B *et al* (9). In brief, dried blood spots (DBS) were self-collected at home. Study participants were provided with material kits including one protein saver card (Whatman 903, Z761575) and two sterile contact-activated lancets (BD microtainer, 366594) for sample collection. Written instructions were provided to study participants. DBS cards were allowed to air dry for shipping in an airtight bag with a desiccant and humidity indicator to maintain sample integrity. Samples were stored at −20 °C upon arrival at the processing facility. This protocol allowed for submission of a series of up to five DBS per participant at each study time point.

### DBS reconstitution

DBS samples were collected by punching four 6 mm spots from saturated sample collection cards using a BSD 300 DBS puncher, as described by McMillan B *et al* (9). In brief, punches were then eluted by adding 350 µL of buffer comprised of dipotassium phosphate buffered saline (PBS) supplemented with 0.5% w/v sodium azide and 1.5% w/v bovine serum albumin (Ortho Clinical Diagnostics). Samples were agitated for 30 seconds by vortex and allowed to soak overnight at 4 °C to allow for reconstitution. Sample eluent was used immediately and the remaining sample was preserved at −20 °C.

### Serological testing and analysis of dried blood spot samples

Processing of DBS samples was carried out as follows: following overnight incubation of DBS punches, 10 µL of the resultant eluate was diluted 1:500 in Diluent 100 provided (Meso Scale Discovery [MSD], R50AA-2). Serological analysis of coronavirus-specific IgG antibody titres was performed as per the manufacturer’s instructions using a V-PLEX COVID-19 Coronavirus Panel 2 (MSD, USA; Cat. no. K15369U), which includes spike antigens of all six tested coronaviruses (NL63, 229E, HKU1, OC43, SARS-CoV-1, and SARS-CoV-2). Change in electrochemiluminescence from baseline was measured in a MESO QuickPlex SQ 120MM instrument (MSD, USA), with resultant values analyzed using machine-compatible DISCOVERY WORKBENCH 4.0 Analysis Software (MSD, USA). The use of this MSD immunoassay has been previously validated for detection of both anti-SARS-CoV-2 and anti-endemic coronavirus IgG (11). All six spike antigens are coated on the plates at the same concentration, allowing for direct comparison between respective targeting antibody titres. RBD and nucleocapsid antigen constructs have a different molecular weight and cannot be directly compared as the number of molecules per spot differs.

Samples were classified as either SARS-CoV-2 seropositive or seronegative based on the presence or absence of detectable levels of SARS-CoV-2 specific antibodies above the predetermined cut-offs (see ‘Serum cross-validation and threshold determination’ below), and/or report of a positive PCR test (8). Samples were classified as: “Negative” in the absence of detectable SARS-CoV-2 specific antibodies above threshold; or “Positive” if samples contained both anti-spike (S) and anti-nucleocapsid (N) antibodies, indicating natural infection with or without vaccine-induced immunity. Individuals with a vaccine response only were confirmed by an anti-S antibody response and negative anti-N response, indicating vaccine-induced immunity only, though it is not possible to fully rule out natural infection followed by anti-N waning.

### Serum cross-validation

DBS testing results were cross-validated by comparing anti-spike protein IgG levels measured in patient-matched serum samples from the ASSESS-DBS study (8). Samples were collected from incarcerated adult individuals or workers in correctional facilities in British Columbia. Demographic variables were not provided by study administrators and are not included in our analysis as samples were used purely for validation purposes.

Patient blood samples were collected into 5 mL tubes (BD vacutainer SST tubes; Cat. no. 367986) by venipuncture performed by trained phlebotomists. Serum was obtained for testing by centrifugation before long term storage at −20 °C. DBS samples were collected in the facility, reconstituted, and stored as described above. MSD analysis of antigen-specific IgG levels was carried out as described above. Matched samples from 174 adult individuals were used in this study.

### Community-level endemic coronavirus circulation

Circulation of endemic coronaviruses 229E, NL63, HKU1, and OC43 was determined throughout the 2019-2020, 2020-2021, and 2021-2022 surveillance years via publicly available Respiratory Virus Detection Surveillance System (RVDSS) ‘FluWatch’ reports (12). Public data on individual endemic circulation was obtained from the BC Centre for Disease Control testing site in Vancouver, British Columbia. Data is presented as percent positivity out of the total number of tests run.

### ACE2 binding and surrogate SARS-CoV-2 neutralization assays

To determine the extent which reconstituted patient DBS samples could neutralize antigens representative of SARS-CoV-2 wildtype and variant strains, we used a V-PLEX COVID-19 Panel 13 (ACE2) Kit (Meso Scale Discovery [MSD], USA; Cat. no. K15466U). This assay includes spike proteins isolated from the index virus (GISAID Accession ID: EPI_ISL_402124) and variants of concern B.1.1.7 (Alpha), B.1.351 (Beta), B.1.526.1 (Iota), B.1.617, B.1.617.1 (Kappa), B.1.617.2 (Delta), B.1.617.3, P.1 (Gamma), and P.2 (Zeta). The assay was carried out as per the manufacturer’s instruction. Change in electrochemiluminescence from baseline, representative of the amount of antigen-bound patient antibodies, was measured in a MESO QuickPlex SQ 120MM instrument (MSD, USA), with resultant values analyzed using machine-compatible DISCOVERY WORKBENCH 4.0 Analysis Software (MSD, USA). Assay output is a measure of labelled-ACE2 secondary antibody directly binding to plate-bound antigen. Therefore, direct output values reflect the amount of unbound target antigen. Values are presented as percent neutralization.

### Statistical analysis and data visualization

Data and statistical analysis was carried out in R (4.1.3) using the packages tidyverse (1.3.1)(13), rstatix (0.7.2)(14), GGally (2.1.2)(15) and Desctools (0.99.48)(16). All antibody concentration measurements were log-transformed prior to analysis. Unless otherwise stated, analysis of normally distributed data was performed using a Welch’s t-test or one-way ANOVA to compare IgG responses across study visits, with Bonferroni correction for multiple comparisons. Aggregate results represent the geometric mean +/− 95% confidence interval, and statistical significance is represented by \**p* < 0.05, \*\**p* < 0.01, \*\*\**p* < 0.001 and \*\*\*\**p* < 0.0001. Spearman correlation analysis between variables was performed in R using the ggpubr (0.6.0)(17) package (*p*- and rho-values representing Spearman correlation values; lines representing linear regression +/− 95% confidence intervals). Antibody terciles were calculated using the fabricatr package (1.0.0)(18). Fisher’s exact tests were run and visualized using ggstatsplot (0.12.0)(19). Multivariate analysis was performed using the gtsummary (1.7.2)(20) package. Data visualization was carried out in R (4.1.3) using the tidyverse (2.0.0)(13), ggplot2 (3.4.2)(21), table1 (1.4.3)(22), and viridis (0.6.3)(23) R packages.

## Results

### Dried Blood Spot (DBS) testing allows for accurate detection of endemic coronavirus-specific antibodies

To screen coronavirus-specific antibody titers across a large number of participants, we used DBS samples as opposed to serum measurements, to allow participant point-of-care sampling and thereby more effectively access and evaluate population immunity. While the diagnostic accuracy of DBS samples has been previously demonstrated for the detection of IgG antibodies to SARS-CoV-2 (8), we sought to establish the suitability of DBS sample testing for the detection of IgG antibody levels against human endemic coronaviruses. To do this, we compared IgG titers specific to each of the four HCoVs (229E, NL63, HKU1, and OC43) across matched participant serum and DBS samples in a cohort of 174 adults from the ASSESS-DBS study, which sampled adults in correctional facilities prior to COVID-19 vaccination (8). By comparing relative serology, we found that total detected IgG antibody titers were significantly lower in DBS samples compared to serum samples from matched individuals, with a 17- to 19-fold difference across HCoV targets (Figure 1A). However, we observed a strong positive correlation (Figure 1B) between matched serum and DBS IgG titers specific to all four HCoVs: 229E (rho = 0.77; *p* = <2.2e-16), NL63 (rho = 0.9; *p* = <2.2e-16), HKU1 (rho = 0.79; *p* = <2.2e-16), and OC43 (rho = 0.81; *p* = <2.2e-16). These data indicate that DBS samples can be used to accurately evaluate trends in antibody-based immune responses, and support the use of DBS sample measurements to evaluate changes to HCoV antibody levels.

**Figure 1.**
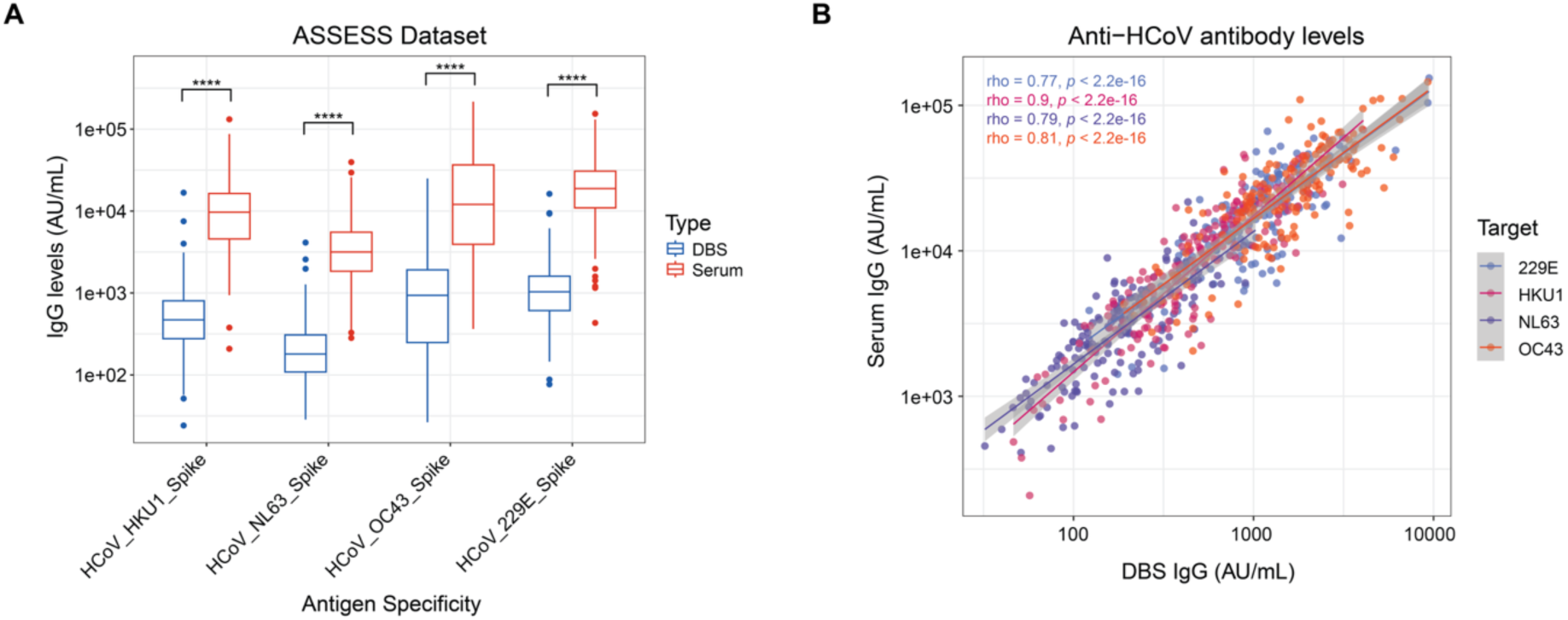
Dried blood spot (DBS) testing allows for robust detection of anti-HCoV antibody levels. A) IgG levels specific to endemic HCoVs 229E, NL63, HKU1 and OC43 as measured in matched DBS and serum samples obtained from the same adult participants. Statistics represent paired t-tests with Bonferroni correction (N = 174). B) Spearman correlation of matched serum and DBS IgG levels across participants. Statistics represent Spearman’s rho; line represents linear regression and confidence interval (N = 174 participants).

### Pre-vaccination levels of anti-HCoV IgG are not related to participant age or sex

Having validated DBS sampling for the evaluation of anti-HCoV humoral immunity, we sought to use DBS testing to investigate the prevalence of HCoV-specific IgG within the PREVENT-COVID study cohort, as described by McMillan B *et al* (9). In brief, a total of 611 individuals aged 50-87 were recruited (median age = 65.3; Table S1), of which 64.5% were female and 35.5% were male, and 64.7% identified as women and 35.1% identified as men. The study population was predominantly white (87.6%), with the next most prevalent self-identified ethnicity as Asian (6.7%). The majority of individuals (65%) identified as having a pre-existing health condition at enrollment, including but not limited to hyper- or hypo-tension, diabetes, or asthma or allergies. At the beginning of the study, almost all individuals were unvaccinated and SARS-CoV-2 seronegative, with only six individuals displaying SARS-CoV-2 seropositivity which were excluded from downstream analysis.

Before investigating temporal patterns in the humoral response to HCoVs we first established baseline differences in IgG specific to Spike proteins of 229E, NL63, HKU1 and OC43. 304 study participants provided samples at visit 1. Of these, samples from 262 individuals were collected the day of their first vaccine dose and were thereby considered ‘baseline’ samples, providing a snapshot of community-level immunity pre-vaccination. All individuals included in baseline analysis were unvaccinated and uninfected, without detectable anti-S and anti-N responses or a positive PCR test. We found significantly different levels of IgG antibodies targeting each of the four HCoVs, with OC43 Spike-specific IgG levels being the highest, and NL63 Spike-specific IgG levels being the lowest (Figure 2A). This relationship was consistent across the collection period and was not influenced by HCoV circulation in the community immediately prior to study enrollment (Figure 2B; Figure S1A). We hypothesized that HCoVs with the highest IgG antibody titers (OC43 and 229E) would have been circulating most recently within the wider population, accounting for their higher baseline antibody abundance. However, we found that all four HCoVs were detected within the population during the preceding 2019-2020 season, with the highest testing positivity rate observed for NL63 and HKU1 (14.75% for both in week 11; Figure S1B).

**Figure 2.**
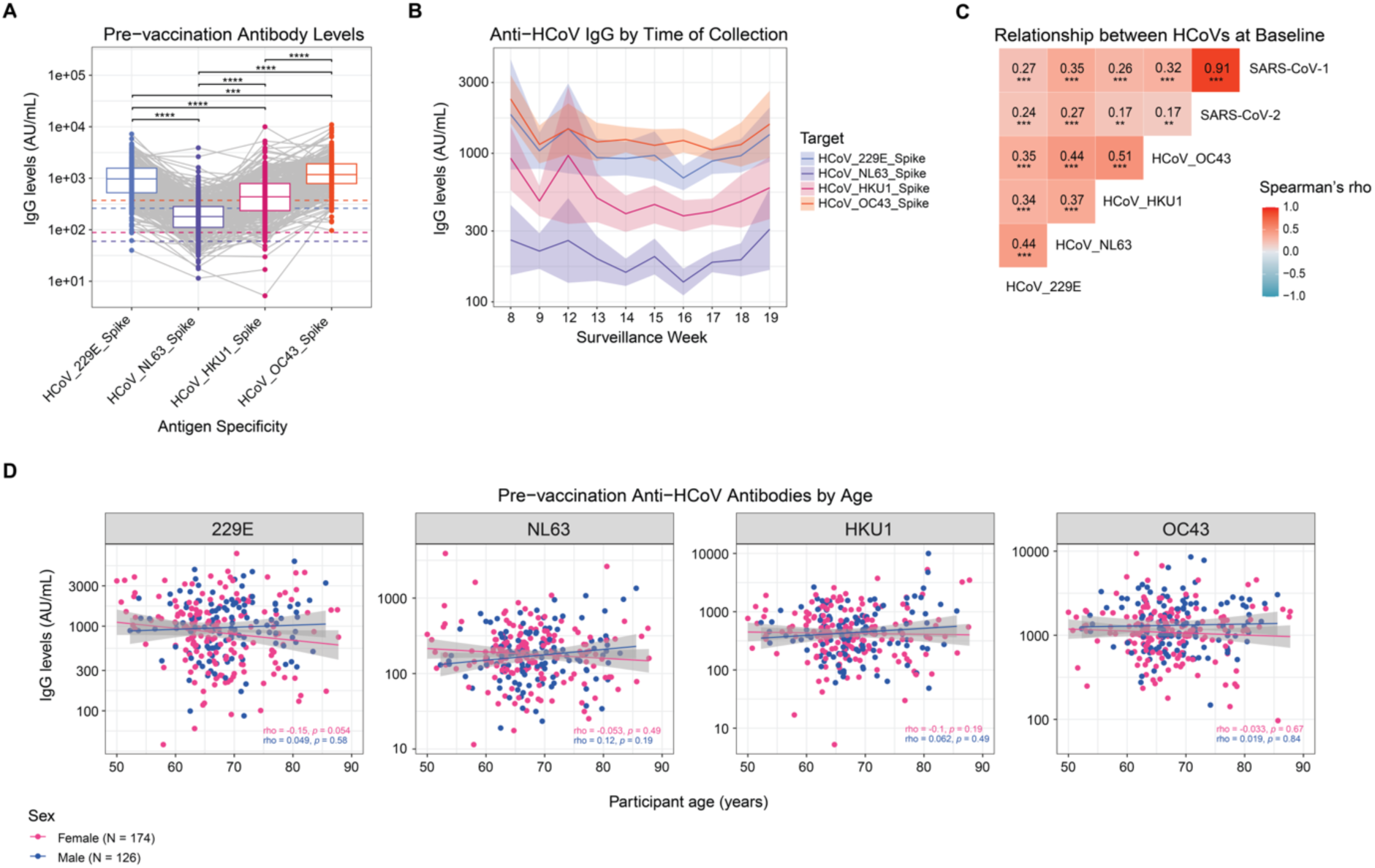
Pre-vaccination levels of anti-HCoV IgG are not related to participant age or sex. A) IgG antibody levels to four endemic coronaviruses (229E, NL63, HKU1, and OC43) at study start, prior to vaccination and/or SARS-CoV-2 infection. Statistics represent paired t-tests with Bonferroni and Holm post-hoc test (N = 304-305). Dotted lines represent individual cut-offs for HCoV seropositivity, coloured by HCoV. B) Anti-HCoV IgG levels by time of collection during the 2020-2021 Flu season (September-August), presented by surveillance week (N = 4-72). Line represents geometric mean +/− 95% confidence interval (CI). Colours represent individual HCoVs. C) Relationship between anti-HCoV IgG, as well as IgG specific to SARS-CoV-1 and SARS-CoV-2 spike proteins at study start (baseline: pre-vaccination/SARS-CoV-2 infection; N = 304). Values represent Spearman’s rho. Colour indicates correlation strength and direction. D) Spearman correlation of pre-vaccination anti-endemic IgG by participant age (N = 174 Female; N = 126 Male). Colours represent participant sex. Values represent Spearman’s rho. Line represents linear regression +/− 95% CI.

Previous studies have demonstrated the cross-reactivity of anti-coronavirus antibodies in binding to more than one viral species (24–27). As such we next wanted to determine how anti-Spike antibodies against the four endemic coronaviruses, as well as SARS-CoV-1 and SARS-CoV-2, correlate at baseline in matched samples. We hypothesized that antibodies targeting the OC43 and HKU1 would correlate most strongly with each other, as well as with SARS-CoV-1 and SARS-CoV-2, which are also beta-coronaviruses. Indeed, we observed that OC43 and HKU1 were more strongly correlated with each other than to alpha-coronaviruses 229E and NL63 (Figure 2C). However, we observed a weak positive correlation (rho = 0.18-0.21) across all HCoVs when compared to anti-SARS-CoV-2 Spike-specific IgG, with the strongest association to NL63 (Figure 2C). To determine the relationship between anti-endemic humoral immunity and older age, we performed correlation analysis of spike-specific IgG for each HCoV by participant age and sex (Figure 2D). We expected anti-HCoV antibody levels to decrease with increasing age, however, we found that baseline antibody levels did not correlate with age in study individuals aged 50-87. We further found no relationship between participant sex and HCoV antibody titers across all four HCoVs. Together, these data characterize the baseline profile of IgG levels which were stable over time of collection and across participant demographics, and which can be used to assess vaccine- and infection-induced changes to anti-HCoV humoral immunity.

### Anti-OC43 and -HKU1 IgG levels increase with COVID-19 vaccination and wane between doses

Having characterized baseline levels of anti-HCoV IgG, we next investigated whether anti-HCoV antibody levels are impacted by COVID-19 vaccination. Further DBS samples were collected from participants before and after the two-dose regimen, as well as following an additional two booster doses (Figure 3A). As per Canadian National Advisory Committee on Immunization (NACI) guidelines, most vaccine doses 1 and 2 were received within an 8-12 week period (82%), with booster doses 1 and 2 following at 6- and 12-months post-dose 2, respectively. To specifically profile vaccine-induced effects on anti-HCoV humoral immunity, we restricted our analysis to only those individuals who remained seronegative for SARS-CoV-2 anti-N throughout the study. Measurement of anti-spike IgG for each HCoV across study visits revealed a significant increase of IgG specific to the two beta-coronaviruses following each vaccine dose, with the greatest response noted in anti-OC43 IgG levels (Figure 3B). Interestingly, anti-betacoronavirus IgG levels demonstrated significant waning between vaccine doses, with anti-HKU1 levels dropping to baseline at 7-months post-booster dose 1. In contrast, we observed no changes in 229E- or NL63-specific IgG titers with vaccination. Circulating HCoV positivity rates in British Columbia Canada further confirm this was unlikely to result from concurrent natural HCoV infection (Figure S2A). These data suggest an interplay between COVID-19 vaccination-induced humoral immune responses targeting coronaviruses.

**Figure 3.**
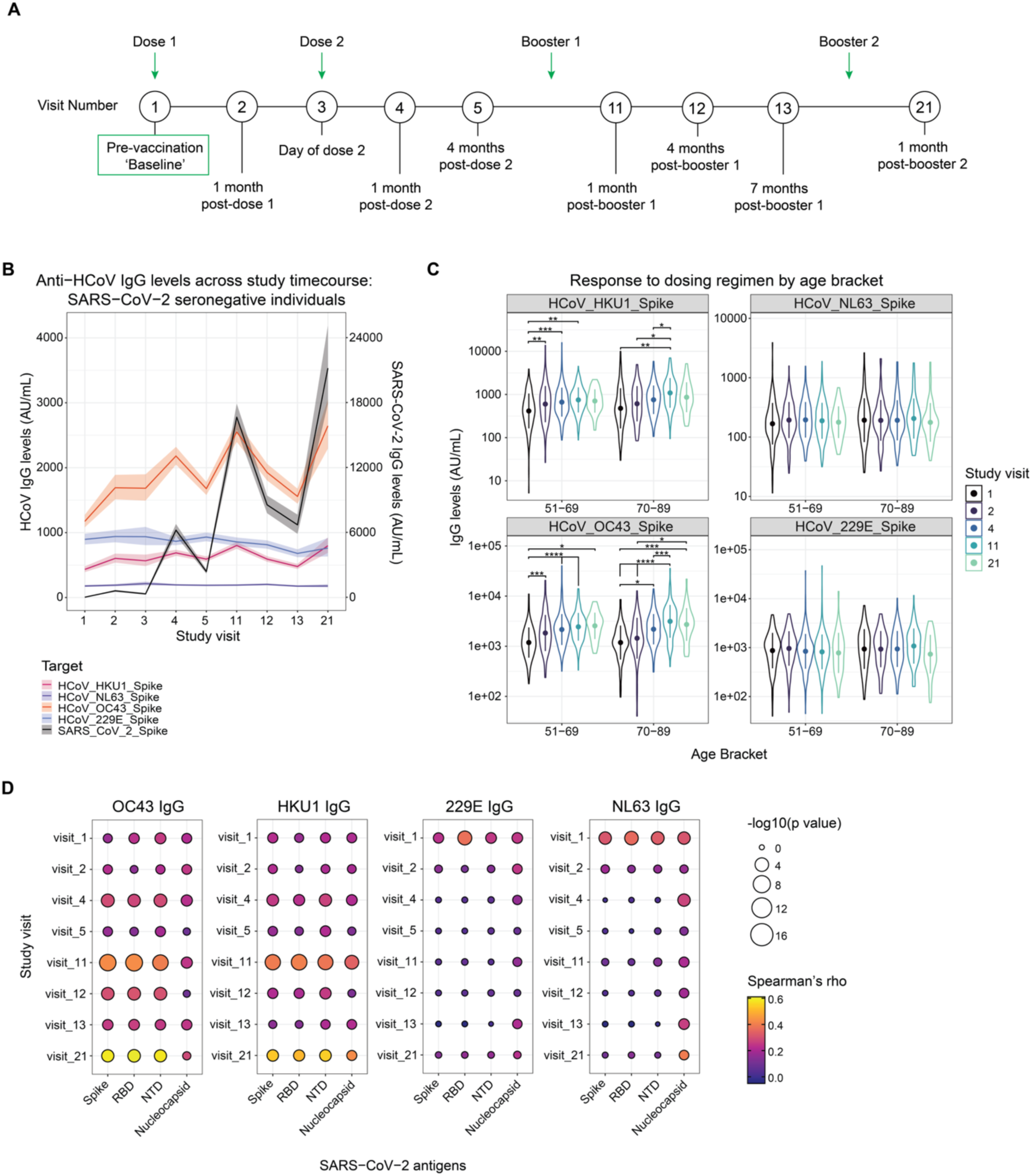
Vaccine-induced humoral immunity to endemic coronaviruses in SARS-CoV-2 seronegative adults. A) Timeline of study visits relative to vaccination as analyzed in this study. Geometric mean and 95% confidence interval (CI) of anti-HCoV and anti-SARS-CoV-2 spike specific IgG levels across study visits (N = 94-505). Colours represent individual coronaviruses. SARS-CoV-2 anti-S antibodies are presented on a 1/6^th^ scale to compare related patterns in lower anti-HCoV IgG levels. C) Anti-endemic anti-S IgG at baseline and 1 month following each vaccine dose (N = 43-378). Data presented by participant age bracket, coloured by study visit. D) Spearman correlation analysis of anti-HCoV antibody titres and antibodies specific to SARS-CoV-2 antigens by study visit (N = 87-489). Size represents -log10(*p-*value). Colour represents Spearman’s rho.

Older individuals are known to mount a weaker SARS-CoV-2 anti-S immune response following COVID-19 vaccination than younger adults (28). Therefore, we next sought to determine whether older age impacted the degree of anti-HCoV boosting following COVID-19 vaccination. To do this, participants were segmented into age brackets as <70 and 70+ years of age to compare antibody titres. No significant differences were observed between age brackets, even when subdivided further into 10-year blocks (Figure 3C; Figure S2B). However, we did note differences in the degree of anti-HCoV antibody boosting post-dose. Anti-HKU1 antibodies were only significantly increased in individuals aged 70+ after three total vaccine doses as compared to after one vaccine dose in individuals <70 (Figure 3C). In contrast, both individuals <70 and 70+ experienced significant increases in anti-OC43 IgG following dose 1 (visit 2) and booster dose 1 (visit 11;Figure 3C). Significant increases in post-vaccination anti-OC43 IgG were primarily driven by individuals aged 61-81 (Figure S2B). No further patterns of anti-alpha-coronavirus IgG were revealed as a result of categorization by age bracket (Figures 3C; S2B). Overall, we observed a large spread of IgG values across all four HCoVs, suggesting that additional individual factors influence antibody titers.

In our study, participants received one of four vaccine types approved by Health Canada at each dose: mRNA vaccines BNT162b2 (Pfizer-BioNTech) or mRNA-1273 (Moderna), or viral vector vaccines ChAdOx1-S (Oxford/AstraZeneca) or Ad26.COV2.S (Janssen). Vaccine types received per dose are summarized in Table S2, with the majority of participants having received the BNT162b2 vaccine. Considering the potential differences in immunogenicity of vaccine types in older adults, we investigated whether the vaccine product received influences anti-HCoV IgG titers, focusing on the IgG response to the initial two-dose regimen. Of individuals with matched values for baseline, 1-month post-dose 1 and 1-month post-dose 2, all of them received either two doses of BNT162b2, two doses of mRNA-1273, or a mix of the two. While mRNA-1273 has been previously shown to induce a stronger SARS-CoV-2 specific humoral immune response, we found no difference in IgG levels specific to HCoVs in individuals aged 60+ (Figure S2C) (29). Furthermore, while a previous study showed that participants who received at least one dose of mRNA-1273 had higher anti-SARS-CoV-2 IgG levels, we did not find that participants who received mRNA-1273 at any point during the study displayed higher anti-endemic antibody titers (Figure S2D) (29).

As anti-OC43 and HKU1 IgG boosts with COVID-19 vaccination, we wanted to further investigate the relationship between levels of anti-endemic IgG and anti-SARS-CoV-2 IgG. We observed a positive correlation between OC43- and HKU1-specific IgG with IgG targeting SARS-CoV-2 spike, receptor-binding domain (RBD), and N-terminal domain (NTD) antigens, which was strongest at visits 11 and 21, representing 1-month post-booster 1 and 1-month post-booster 2, respectively (Figure 3D). As the Health Canada-approved COVID-19 vaccines contain only S-protein antigen and not SARS-CoV-2 nucleocapsid (N), we did not expect the relationship between HCoV-specific SARS-CoV-2 anti-N antibodies to change post-vaccination. However, we observed a moderate positive correlation between anti-N IgG and HKU1 and NL63 antibody titers at visit 21 (rho = 0.427 and rho = 0.397, respectively). HCoVs 229E and NL63 did not correlate with SARS-CoV-2 spike-specific antibodies post-vaccination, despite having the strongest positive relationship at baseline. Taken together, these data suggest that humoral responses to endemic beta-coronaviruses are induced by COVID-19 vaccination in the absence of natural circulating HCoV infection.

### SARS-CoV-2 infection-induced immunity impacts overall anti-endemic IgG levels but not foldchange post-vaccination

Until this point all analyses were completed in SARS-CoV-2 negative individuals. We next sought to evaluate the effect of natural SARS-CoV-2 infection on anti-HCoV antibody levels. To do this we profiled endemic anti-S antibodies across the study time course in SARS-CoV-2 seropositive individuals, as identified by their combined anti-S and anti-N serology or a positive PCR test result. As study participants became infected at different times throughout the study, we chose to include only post-infection serology measurements in our analysis, with the exception of baseline (visit 1) values. We noted an overall increase in OC43- and HKU1-specific IgG levels across all study time points in SARS-CoV-2 seropositive individuals (Figure 4A), as compared to seronegative values. There was no detected influence of SARS-CoV-2 infection on anti-NL63 responses across visits.

**Figure 4.**
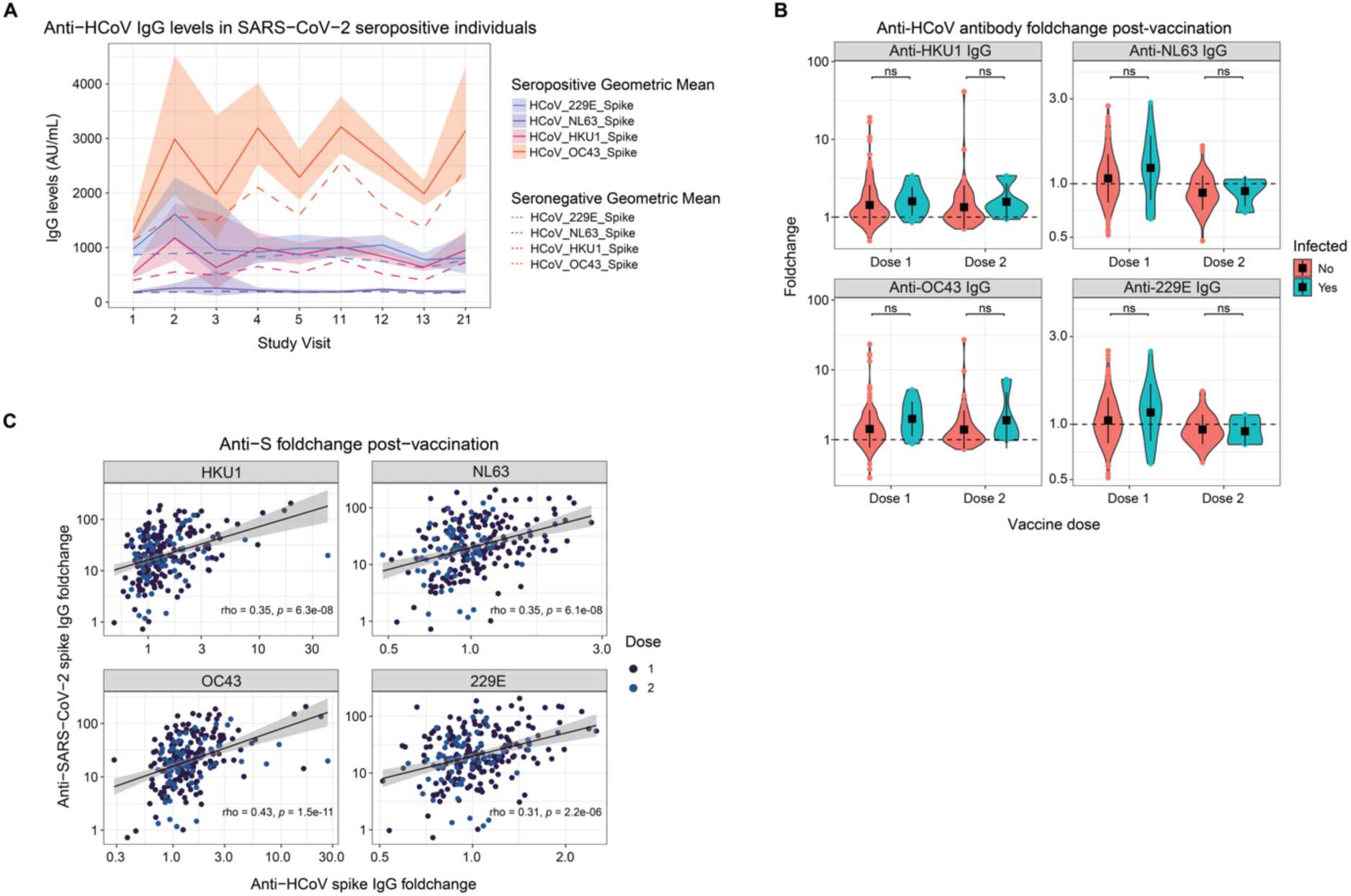
COVID-19 infection-induced humoral response to endemic coronaviruses. A) Geometric mean and 95% confidence interval (CI) of anti-HCoV IgG levels across study visits. Includes post-infection values only, with the exception of visit 1/baseline (N = 4-130). Colours represent individual HCoVs. Solid line and ribbon represents geometric mean ± 95% CI across SARS-CoV-2 seropositive individuals. Dashed line represents geometric mean across SARS-CoV-2 seronegative individuals for reference. B) Comparison of vaccine- and infection-induced foldchange in IgG antibody levels specific to each HCoV (N = 4-253). Colours represent SARS-CoV-2 infection status. C) Spearman correlation analysis of anti-HCoV antibody foldchange and SARS-CoV-2 anti-S foldchange post-vaccination with the two-dose regimen (N = 93-270). Colour represents vaccine dose. Values represent Spearman’s rho. Line represents linear regression +/− 95% CI.

Despite significant differences in OC43 and HKU1 humoral responses, we noted a much greater spread of values across visits following SARS-CoV-2 infection. We did not find any association between participant sex and vaccine-induced IgG titers in SARS-CoV-2 seropositive individuals (Figure S3A), although individuals age 70+ demonstrated a trend towards increased anti-HCoV responses post-infection which was significant at select time points post-dose 2 (Figure S3B). The remaining spread of IgG values was likely due to the difference in timing of infection and subsequent waning of post-infection boosts to anti-HCoV immunity across individuals. To further investigate differences in vaccine-induced and infection-induced immunity, we compared the foldchange in anti-HCoV antibodies pre- and post-doses 1 and 2 in vaccinated seronegative and seropositive individuals (Figure 4B). Despite the overall increase in antibody levels against HKU1 and OC43 in SARS-CoV-2 infected individuals, there was no significant difference in antibody foldchange post-vaccination between SARS-CoV-2 infection positive and negative individuals.

Of note, some individuals experienced a negative foldchange in anti-HCoV antibody levels post-vaccination, with up to 68.42% experiencing a decreased foldchange pre- and post-dose 1 and dose 2 to 229E and NL63 antigen targets (Figure 4B). In comparison, almost all individuals experienced an anti-SARS-CoV-2 spike protein IgG foldchange >1 post-vaccination at either dose 1 or dose 2 (Figure S3C). We did not find a relationship between any one demographic factor and whether participants experienced increased anti-HCoV antibodies post-vaccination or not (data not shown), although we found that individuals with higher baseline HKU1 antibody levels generally demonstrated a lower foldchange post-vaccination regardless of age (Figure S3D; *p* = <0.001).

We found that post-vaccination foldchange in anti-HCoV spike-specific antibody levels were positively correlated with post-vaccination foldchange in anti-SARS-CoV-2 anti-S antibody levels, including the two alpha-coronaviruses NL63 and 229E (Figure 4C). A strong positive relationship was again observed between overall antibody titres of anti-SARS-CoV-2 IgG recognizing the Spike, RBD, and NTD, and antibody levels against HKU1 and OC43 (Figure S3E), with a stronger correlation in vaccinated SARS-CoV-2-infected individuals as compared with infection-naïve individuals. IgG specific for NL63 again correlated most strongly with SARS-CoV-2 anti-N IgG levels (Figure S3E). Together, these data suggest an important relationship between anti-HCoV antibody levels and humoral immune responses to SARS-CoV-2 antigen exposure across all participants regardless of age or other demographic factor.

### Humoral responses to endemic coronaviruses are associated with decreased SARS-CoV-2 anti-N waning and increased surrogate neutralization of SARS-CoV-2 variants

Considering the positive correlation between increased SARS-CoV-2 anti-S and anti-HCoV IgG levels post-vaccination (Figure 4C), and observed waning of anti-OC43 and anti-HKU1 antibodies at >1 month following each COVID-19 vaccination (Figures 3B and 4A), we next investigated whether anti-HCoV antibody levels were further associated with waning of protective anti-SARS-CoV-2 humoral responses in between vaccine doses. To compare waning of anti-SARS-CoV-2 and anti-HCoV antibody titers, we calculated the foldchange per month post-vaccination. While waning of SARS-CoV-2 IgG occurs more rapidly than anti-HCoV IgG, we observed a strong positive correlation between SARS-CoV-2 anti-S waning post-vaccination and waning of IgG specific to the spike proteins of all four endemic coronaviruses in both SARS-CoV-2 infected and infection-naïve participants (Figure S4A). This relationship was independent of the HCoV-specific antibody levels at the time of vaccination (Figure S4B).

As waning of SARS-CoV-2 anti-N antibodies is associated with increased susceptibility to SARS-CoV-2 re-infection (30), we further compared anti-HCoV antibody levels to anti-N specific IgG over time in previously infected individuals. To do this we determined the foldchange in anti-N antibody levels over a period of three months post-SARS-CoV-2 infection, during which selected individuals were neither vaccinated nor re-infected (the period between detected infection at one and four months post-booster 1 [visits 11 and 12, respectively]). Interestingly, we found that individuals with the highest levels of anti-NL63 antibodies at the time of first detected infection demonstrated a significantly lower degree of waning post-infection (Figure S4C). These data align with our previous finding that NL63 IgG levels correlate the most strongly of all endemic coronaviruses with the SARS-CoV-2 anti-N response (Figures 3D and S3E). Indeed, we identified a trend towards decreased anti-N seroreversion in individuals with high tercile NL63-specific IgG levels at baseline, with only 17% of individuals in the high tercile group experiencing seroreversion post-infection, as compared to 38% in the low tercile group (Figure S4D).

Considering the association of anti-HCoV IgG levels with immune waning, we investigated HCoV antibody levels as a risk factor for SARS-CoV-2 infection. We wanted to determine whether individuals who had a positive or negative anti-HCoV IgG foldchange post-vaccine dose 1 or dose 2 were more or less likely to become infected with SARS-CoV-2 over the course of the study. We performed multivariate analysis of subsequent infection risk by both baseline HCoV antibody tercile and post-vaccination HCoV antibody foldchange, accounting for participant age, sex, pre-existing medical conditions and BMI. However, we did not find any significant relationship between infection, anti-HCoV responses, or any of the demographic variables investigated (Tables S3 and S4; data not shown). Furthermore, we did not find that age, sex, pre-existing medical condition or BMI significantly increased the odds of having a high tercile anti-HCoV IgG response at study start, although there was a trend towards an increased odds ratio of high tercile baseline NL63 antibodies in individuals aged 70+, as well as a trend towards high baseline OC43 levels in individuals with pre-existing conditions (Table S5).

To functionally investigate the relationship between anti-HCoV responses and SARS-CoV-2 susceptibility, we next investigated whether HCoV-specific antibody levels were associated with SARS-CoV-2 neutralization. To do this, we used a surrogate neutralization assay in which reconstituted participant DBS samples were tested for their ability to block the binding of angiotensin-converting enzyme 2 (ACE2) to the SARS-CoV-2 spike protein. We found that percent neutralization of wild-type SARS-CoV-2 positively correlated with IgG levels specific for the two beta-coronaviruses OC43 and HKU1 in both SARS-CoV-2 infected and infection-naïve participants, with no significant differences between age brackets (Figure 5A-D). This relationship was consistent across nine additional variants of concern (B.1.1.7, B.1.351, B.1.526.1, B.1.617, B.1.617.1, B.1.617.2, B.1.617.3, P.1, and P.2) despite temporal differences in their circulation within the study population (Table S6). Furthermore, participant age and sex did not impact the participant’s ability to block ACE2 binding (Figure 5A-D; data not shown). Taken together, humoral immune responses targeting endemic coronaviruses may be indicative of SARS-CoV-2 neutralization and overall host susceptibility to SARS-CoV-2 infection.

**Figure 5.**
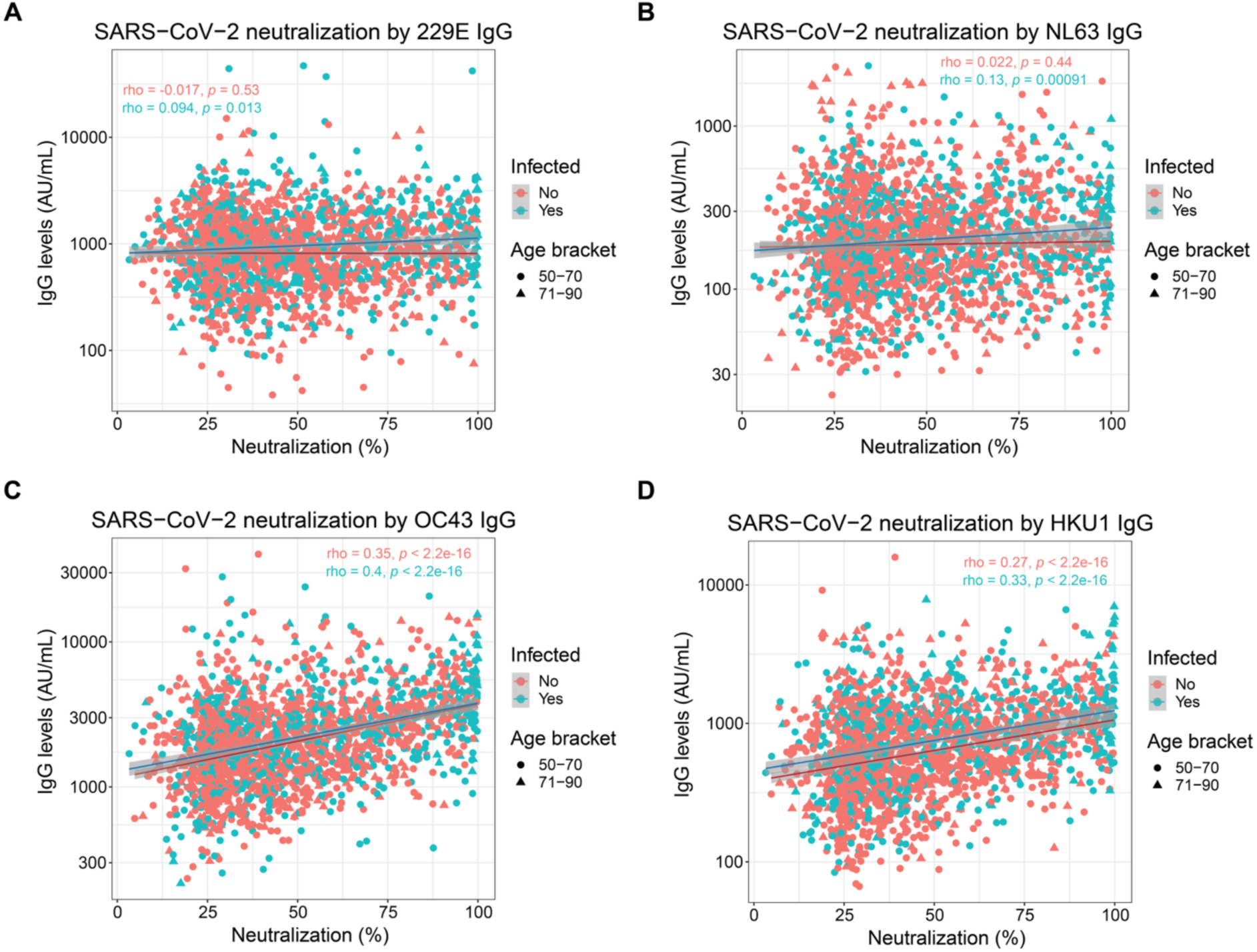
Surrogate SARS-CoV-2 neutralization by anti-endemic IgG levels. A-D) Surrogate neutralization of wild-type SARS-CoV-2 by anti-endemic IgG titres. Statistics represent Spearman correlation analysis. Colour represents SARS-CoV-2 seropositivity. Symbol shape represents participant age bracket. Values represent Spearman’s rho. Line represents linear regression +/− 95% CI.

## Discussion

Most individuals over the age of 10 demonstrate robust humoral immunity against the four circulating endemic coronaviruses (229E, NL63, HKU1, and OC43), with estimated re-infection every two to three years (2,3). It follows that by the age of 50, older adults have received significant exposure to coronaviruses which could impact immune responses to subsequent infection, either with HCoVs or related pandemic coronaviruses. In this study, we assessed the relationship between IgG antibodies specific to HCoVs and to SARS-CoV-2 within a prospective cohort of 611 community-dwelling older adults in British Columbia, Canada.

Analysis of pre-vaccination HCoV IgG levels in SARS-CoV-2 seronegative individuals allowed us to establish the humoral immune landscape before SARS-CoV-2 antigen exposure. Baseline responses were characterized by high relative anti-OC43 antibodies and low anti-NL63 antibodies (Figure 2), despite recent circulation of NL63 in the surveillance year prior to pandemic onset (Figure S1). This pattern has been previously observed across infants (< 6 months of age) and non-senior adults (mean age 42.4) (27). While it is possible that HKU1 and NL63 are less immunogenic than OC43 and 229E, the positive relationship between alpha- and between beta-coronavirus antibodies could indicate possible cross-reactivity. For example, between anti-OC43 and anti-HKU1 beta-coronavirus IgG responses, as well as between anti-NL63 and anti-229E alpha-coronavirus IgG responses (Figure 2C). Alternatively, as NL63 and HKU1 recently emerged in humans (in 2004 and 2005, respectively), it may also be that infection by these HCoVs results in immune back-boosting of IgG specific to closely related coronaviruses, rather than full induction of NL63- or HKU1-specific antibody responses (1).

This potential for ‘immune imprinting’ could further explain the increase in OC43 and HKU1 antibody levels resulting from COVID-19 vaccination (Figure 3). This phenomenon has already been reported in the context of COVID-19, whereby an individual’s first antigen exposure, through vaccination or natural infection, determines how an individual responds to subsequent infection by new variant strains (31,32). It is unclear whether the observed boosting of beta-coronavirus specific IgG titres is due to immune back-boosting as a result of immune imprinting, or the result of cross-reactivity of SARS-CoV-2 targeting antibodies towards related coronaviruses. While the SARS-CoV-2 spike protein shares only 25-30% homology to HCoVs (33,34), induction of anti-HCoV immune responses has been observed following COVID-19 vaccination of humans and HCoV-naïve macaques, suggesting possible cross-reactivity of SARS-CoV-2 specific antibodies to endemic coronaviruses (4). Regardless of outcome, anti-HCoV IgG boosting could affect population-wide protection against beta-coronaviruses and possibly lead to reduced rates of OC43 and HKU1 infection and/or domination of HCoV infections by unaffected alpha-coronaviruses. However, due to considerable waning of anti-HCoV responses in between vaccine doses, it is likely that variable time of infection may mask any short-lived protective effects in population-wide screens.

Beta-coronavirus IgG is not only boosted by vaccination but generally exhibits a robust positive relationship with anti-SARS-CoV-2 antibody levels specific to spike, RBD and NTD antigens which is strengthened by repeated COVID-19 vaccination (Figures 3 and 4). Indeed, foldchange of all four HCoVs positively correlated with SARS-CoV-2 anti-S foldchange in the context of both post-vaccination boosting and subsequent waning (Figures 4 and S4). This may indicate that humoral immunity toward HCoVs may also be associated with protection against SARS-CoV-2 infection. Indeed, we found that individuals with higher OC43 IgG levels demonstrated an increased percent neutralization of both the index virus and nine additional circulating variants, as determined by the ability of antibodies to bind to and block ACE2 (Figure 5; Table S6). Due to the parallel relationship between SARS-CoV-2 and HCoV antibodies, it is challenging to determine whether this is simply due to higher corresponding anti-SARS-CoV-2 IgG present in participant samples, rather than a protective effect of high anti-HCoV IgG.

However, we did observe a relationship between NL63-specific IgG levels and both anti-N titres and anti-N waning post-infection, despite a lack of boosting of NL63 following either SARS-CoV-2 vaccination or infection, wherein higher NL63 IgG levels were associated with reduced anti-N waning and a trend towards decreased incidence of anti-N seroreversion during the study time course (Figure S4). Additional studies have also linked NL63 IgG levels to increased protection against SARS-CoV-2 infection in mice and humans (35,36). Of note, NL63 is the only HCoV known to use the ACE2 receptor for human cell entry, as do SARS-CoV-1 and SARS-CoV-2, making it possible that epitope shape influences these interactions (37). Overall, the recent display of common cold symptoms has been shown to decrease the odds of SARS-CoV-2 infection, suggesting a potential protective effect of prior HCoV exposure and antibody responses (25).

Altogether, this study provides a comprehensive longitudinal profile of anti-HCoV IgG responses in community-dwelling older adults. We make key observations about the dynamics of anti-HCoV IgG over the course of repeated vaccination, and consequently highlight the potential for widespread off-target effects of COVID-19 vaccination on humoral immunity which may affect the general circulation of alpha- and beta-coronaviruses. Furthermore, recent studies have demonstrated the potential for CD4^+^ T cells from pre-pandemic samples to recognize epitopes from OC43, NL63 and SARS-CoV-2 and contribute to host protection (38). Such broad population-wide adaptive immune crosstalk in coronavirus responses could drive the evolution of SARS-CoV-2 towards endemic status. A key next step will be to determine whether the observed vaccine-induced effects are the result of immune back-boosting, which could imply a potential for immune blunting following repeated related antigen exposure, or instead due to cross-reactivity of SARS-CoV-2 induced antibodies. Either outcome has implications for antibody responses not only to related HCoVs, but also to wildtype SARS-CoV-2 and emerging variant strains. Further understanding could likewise inform the design of pan-coronavirus vaccines by prioritizing antigens which are least cross-reactive.

## Acknowledgments

The authors would like to thank participants of both the ASSESS-DBS and PREVENT-COVID studies, as well as the dedicated team at the Vaccine Evaluation Center who recruited and screened participants and at the BC Centre for Disease Control Public Health Laboratory who supported sample testing. Thank you to Danielle Luk for helping to process DBS samples from PREVENT-COVID and to Lin Chang for help in accessing FluWatch surveillance reports. This work was funded by grants to MS and ANJ from the Public Health Agency of Canada; ANJ was supported via the Vaccine Surveillance Reference Group and the COVID-19 Immunity Task Force (2122-HQ-000218); MS was supported by the BC Immunization Committee (AWD-018041). BM was supported by a Canadian Immunization Research Network Doctoral Award. GNG was supported by a BC Children’s Hospital Bertram Hofmeister Postdoctoral Fellowship Award and Canadian Immunization Research Network Postdoctoral Fellowship Award.

## Data Availability Statement

Datasets generated and/or analyzed in this manuscript are available, devoid of identifying information, from the corresponding authors upon request.

## Conflict of Interest Statement

MS has been involved in projects funded by GlaxoSmithKline, Merck, Moderna, Pfizer, Sanofi-Pasteur, Seqirus, Symvivo, and VBI Vaccines, with all funds paid to the institute and none received personally. SRB has spoken for and participated in medical advisory board programs with Gilead Sciences Canada, Inc. and AbbVie Corp. Canada. No personal payments were accepted. SRB further received investigator-initiated, unrestricted grant funding from Gilead Sciences Canada, Inc. and AbbVie Corp. Canada via their institution. This funding was unrelated to the current work.

## Notes

### Author Declarations

Ethics approval for the 'PRospEctiVe EvaluatioN of immunity after COVID-19 vaccines' (PREVENT-COVID) study was provided by the University of British Columbia Clinical Research Ethics Board (Ref: H20-03951). Written informed consent was obtained from all study participants. Further data was used from the Accelerating SARS-CoV-2 Seroprevalence SurveyS through Dried Blood Spots (ASSESS-DBS) study. Ethics approval for the ASSESS-DBS study was granted by the University of British Columbia Clinical Research Ethics Board (Ref: H20-02402).

## References

1. Corman VM, Muth D, Niemeyer D, Drosten C. Hosts and Sources of Endemic Human Coronaviruses. Adv Virus Res. 2018;100:163–88.

2. Tanunliong G, Liu AC, Kaweski S, Irvine M, Reyes RC, Purych D, et al. Age-Associated Seroprevalence of Coronavirus Antibodies: Population-Based Serosurveys in 2013 and 2020, British Columbia, Canada. Front Immunol. 2022;13:836449.

3. Petrie JG, Bazzi LA, McDermott AB, Follmann D, Esposito D, Hatcher C, et al. Coronavirus Occurrence in the Household Influenza Vaccine Evaluation (HIVE) Cohort of Michigan Households: Reinfection Frequency and Serologic Responses to Seasonal and Severe Acute Respiratory Syndrome Coronaviruses. The Journal of Infectious Diseases. 2021 Jul 1;224(1):49–59.

4. Grobben M, van der Straten K, Brouwer PJ, Brinkkemper M, Maisonnasse P, Dereuddre-Bosquet N, et al. Cross-reactive antibodies after SARS-CoV-2 infection and vaccination. Elife. 2021 Nov 23;10:e70330.

5. Asamoah-Boaheng M, Grunau B, Karim ME, Jassem AN, Bolster J, Marquez AC, et al. Are higher antibody levels against seasonal human coronaviruses associated with a more robust humoral immune response after SARS-CoV-2 vaccination? Front Immunol. 2022;13:954093.

6. Ng KW, Faulkner N, Cornish GH, Rosa A, Harvey R, Hussain S, et al. Preexisting and de novo humoral immunity to SARS-CoV-2 in humans. Science. 2020 Dec 11;370(6522):1339–43.

7. Pinotti F, Wikramaratna PS, Obolski U, Paton RS, Damineli DSC, Alcantara LCJ, et al. Potential impact of individual exposure histories to endemic human coronaviruses on age-dependent severity of COVID-19. BMC Med. 2021 Jan 12;19:19.

8. Nikiforuk AM, McMillan B, Bartlett SR, Márquez AC, Pidduck T, Kustra J, et al. Performance of Immunoglobulin G Serology on Finger Prick Capillary Dried Blood Spot Samples to Detect a SARS-CoV-2 Antibody Response. Microbiol Spectr. 2022 Apr 27;10(2):e0140521.

9. McMillan B, Gaultier GN, Marquez AC, Valadbeigy T, So B, Schwartz S, et al. 1165. Longitudinal Evaluation of SARS-CoV-2 Antibody Response Using Dried Blood Spot Samples Following Vaccination with Three and Four Doses of mRNA-1273, BNT162b2 and/or ChAdOx1-S in Adults Aged 50 and Above: Interim Analysis from the PREVENT-COVID Study. Open Forum Infect Dis. 2023 Nov 27;10(Suppl 2):ofad500.1005.

10. Hogan CA, Jassem AN, Sbihi H, Joffres Y, Tyson JR, Noftall K, et al. Rapid Increase in SARS-CoV-2 P.1 Lineage Leading to Codominance with B.1.1.7 Lineage, British Columbia, Canada, January–April 2021 - Volume 27, Number 11—November 2021 - Emerging Infectious Diseases journal - CDC. [cited 2025 Jul 9]; Available from: https://wwwnc.cdc.gov/eid/article/27/11/21-1190_article

11. Li FF, Liu A, Gibbs E, Tanunliong G, Marquez AC, Gantt S, et al. A novel multiplex electrochemiluminescent immunoassay for detection and quantification of anti-SARS-CoV-2 IgG and anti-seasonal endemic human coronavirus IgG. J Clin Virol. 2022 Jan;146:105050.

12. Canada PHA of. Respiratory virus detections in Canada [Internet]. 2012 [cited 2023 Dec 31]. Available from: https://www.canada.ca/en/public-health/services/surveillance/respiratory-virus-detections-canada.html

13. Wickham H, Averick M, Bryan J, Chang W, McGowan LD, François R, et al. Welcome to the Tidyverse. Journal of Open Source Software. 2019 Nov 21;4(43):1686.

14. Kassambara A. rstatix: Pipe-Friendly Framework for Basic Statistical Tests [Internet]. 2023 [cited 2023 Dec 19]. Available from: https://cran.r-project.org/web/packages/rstatix/index.html

15. Schloerke B, Cook D, Larmarange J, Briatte F, Marbach M, Thoen E, et al. GGally: Extension to “ggplot2” [Internet]. 2023 [cited 2023 Dec 29]. Available from: https://cran.r-project.org/web/packages/GGally/index.html

16. Signorell A, Aho K, Alfons A, Anderegg N, Aragon T, Arachchige C, et al. DescTools: Tools for Descriptive Statistics [Internet]. 2023 [cited 2023 Dec 19]. Available from: https://cran.r-project.org/web/packages/DescTools/index.html

17. Kassambara A. ggpubr: “ggplot2” Based Publication Ready Plots [Internet]. 2023 [cited 2023 Dec 19]. Available from: https://cran.r-project.org/web/packages/ggpubr/index.html

18. Blair G, Cooper J, Coppock A, Humphreys M, Rudkin A, Fultz N, et al. fabricatr: Imagine Your Data Before You Collect It [Internet]. 2022 [cited 2023 Dec 19]. Available from: https://cran.r-project.org/web/packages/fabricatr/index.html

19. Patil I. Visualizations with statistical details: The “ggstatsplot” approach. Journal of Open Source Software. 2021 May 25;6(61):3167.

20. Sjoberg DD, Whiting K, Curry M, Lavery JA, Larmarange J. Reproducible Summary Tables with the gtsummary Package. The R Journal. 2021;13(1):570–80.

21. Wickham H. Modelling for Visualisation. In: Wickham H, editor. ggplot2: Elegant Graphics for Data Analysis [Internet]. Cham: Springer International Publishing; 2016 [cited 2022 Feb 2]. p. 221–40. (Use R!). Available from: 10.1007/978-3-319-24277-4_11

22. Rich B. table1: Tables of Descriptive Statistics in HTML [Internet]. 2023 [cited 2023 Dec 19]. Available from: https://cran.r-project.org/web/packages/table1/index.html

23. Garnier S, Ross N, Rudis B, Sciaini M, Camargo AP, Scherer C. viridis: Colorblind-Friendly Color Maps for R [Internet]. 2023 [cited 2023 Dec 19]. Available from: https://cran.r-project.org/web/packages/viridis/index.html

24. Jaago M, Rähni A, Pupina N, Pihlak A, Sadam H, Tuvikene J, et al. Differential patterns of cross-reactive antibody response against SARS-CoV-2 spike protein detected for chronically ill and healthy COVID-19 naïve individuals. Sci Rep. 2022 Oct 7;12:16817.

25. Aran D, Beachler DC, Lanes S, Overhage JM. Prior presumed coronavirus infection reduces COVID-19 risk: A cohort study. J Infect. 2020 Dec;81(6):923–30.

26. Sagar M, Reifler K, Rossi M, Miller NS, Sinha P, White LF, et al. Recent endemic coronavirus infection is associated with less-severe COVID-19. J Clin Invest. 131(1):e143380.

27. Majdoubi A, Michalski C, O’Connell SE, Dada S, Narpala S, Gelinas J, et al. A majority of uninfected adults show preexisting antibody reactivity against SARS-CoV-2. JCI Insight. 6(8):e146316.

28. San Román J, Candel FJ, Sanz JC, López P, Menéndez-Colino R, Barreiro P, et al. Humoral and Cellular Response after mRNA Vaccination in Nursing Homes: Influence of Age and of History of COVID-19. Vaccines (Basel). 2022 Mar 2;10(3):383.

29. Walmsley SL, Szadkowski L, Wouters B, Clarke R, Colwill K, Rochon P, et al. COVID-19 vaccine antibody responses in community-dwelling adults to 48 weeks post primary vaccine series. iScience. 2023 Apr 21;26(4):106506.

30. Cohen D, Izak M, Stoyanov E, Mandelboim M, Perlman S, Amir Y, et al. Predictors of reinfection with pre-Omicron and Omicron variants of concern among individuals who recovered from COVID-19 in the first year of the pandemic. International Journal of Infectious Diseases. 2023 Jul 1;132:72–9.

31. Reynolds CJ, Gibbons JM, Pade C, Lin KM, Sandoval DM, Pieper F, et al. Heterologous infection and vaccination shapes immunity against SARS-CoV-2 variants. Science. 2022 Jan 14;375(6577):183–92.

32. Reynolds CJ, Pade C, Gibbons JM, Otter AD, Lin KM, Muñoz Sandoval D, et al. Immune boosting by B.1.1.529 (Omicron) depends on previous SARS-CoV-2 exposure. Science. 2022 Jun 14;377(6603):eabq1841.

33. Ratcliffe H, Tiley KS, Longet S, Tonry C, Roarty C, Watson C, et al. Serum HCoV-spike specific antibodies do not protect against subsequent SARS-CoV-2 infection in children and adolescents. iScience. 2023 Nov 21;26(12):108500.

34. Barnes CO, West AP, Huey-Tubman KE, Hoffmann MAG, Sharaf NG, Hoffman PR, et al. Structures of Human Antibodies Bound to SARS-CoV-2 Spike Reveal Common Epitopes and Recurrent Features of Antibodies. Cell. 2020 Aug 20;182(4):828–842.e16.

35. Lesmes-Rodríguez LC, Lambarey H, Chetram A, Riou C, Wilkinson RJ, Joyimbana W, et al. Previous exposure to common coronavirus HCoV-NL63 is associated with reduced COVID-19 severity in patients from Cape Town, South Africa. Frontiers in Virology [Internet]. 2023 [cited 2023 Dec 19];3. Available from: https://www.frontiersin.org/articles/10.3389/fviro.2023.1125448

36. Liu D, Chen C, Chen D, Zhu A, Li F, Zhuang Z, et al. Mouse models susceptible to HCoV-229E and HCoV-NL63 and cross protection from challenge with SARS-CoV-2. Proceedings of the National Academy of Sciences. 2023 Jan 24;120(4):e2202820120.

37. Shirbhate E, Pandey J, Patel VK, Kamal M, Jawaid T, Gorain B, et al. Understanding the role of ACE-2 receptor in pathogenesis of COVID-19 disease: a potential approach for therapeutic intervention. Pharmacol Rep. 2021;73(6):1539–50.

38. Tarke A, Zhang Y, Methot N, Narowski TM, Phillips E, Mallal S, et al. Targets and cross-reactivity of human T cell recognition of common cold coronaviruses. Cell Rep Med. 2023 Jun 20;4(6):101088.

