## Supplementary material for "Humoral immunity to endemic coronaviruses in older adults increases post-COVID-19 vaccination and in correlation with their anti-SARS-CoV-2 responses": Table S1

Supplemental Figures and Tables

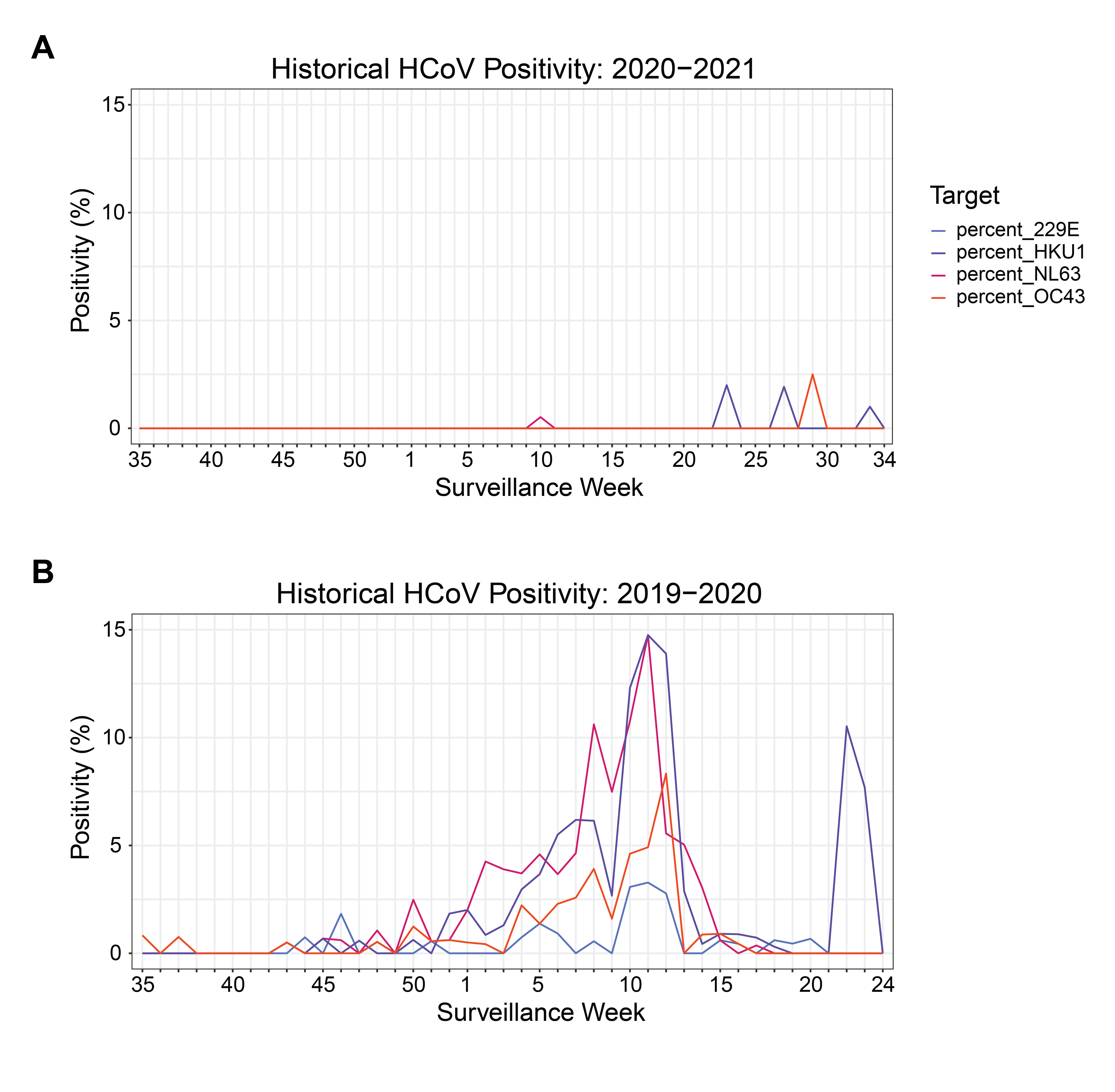

**Figure S1. Endemic coronavirus positivity rates by testing prior to study start and during baseline sampling.** A) HCoV percent positivity rates by surveillance week during the September 2020 – August 2021 season. Colours represent individual HCoVs. B) HCoV percent positivity rates by surveillance week during the September 2019 – August 2020 season. Colours represent individual HCoVs.

**Table S1. PREVENT-COVID study cohort.**

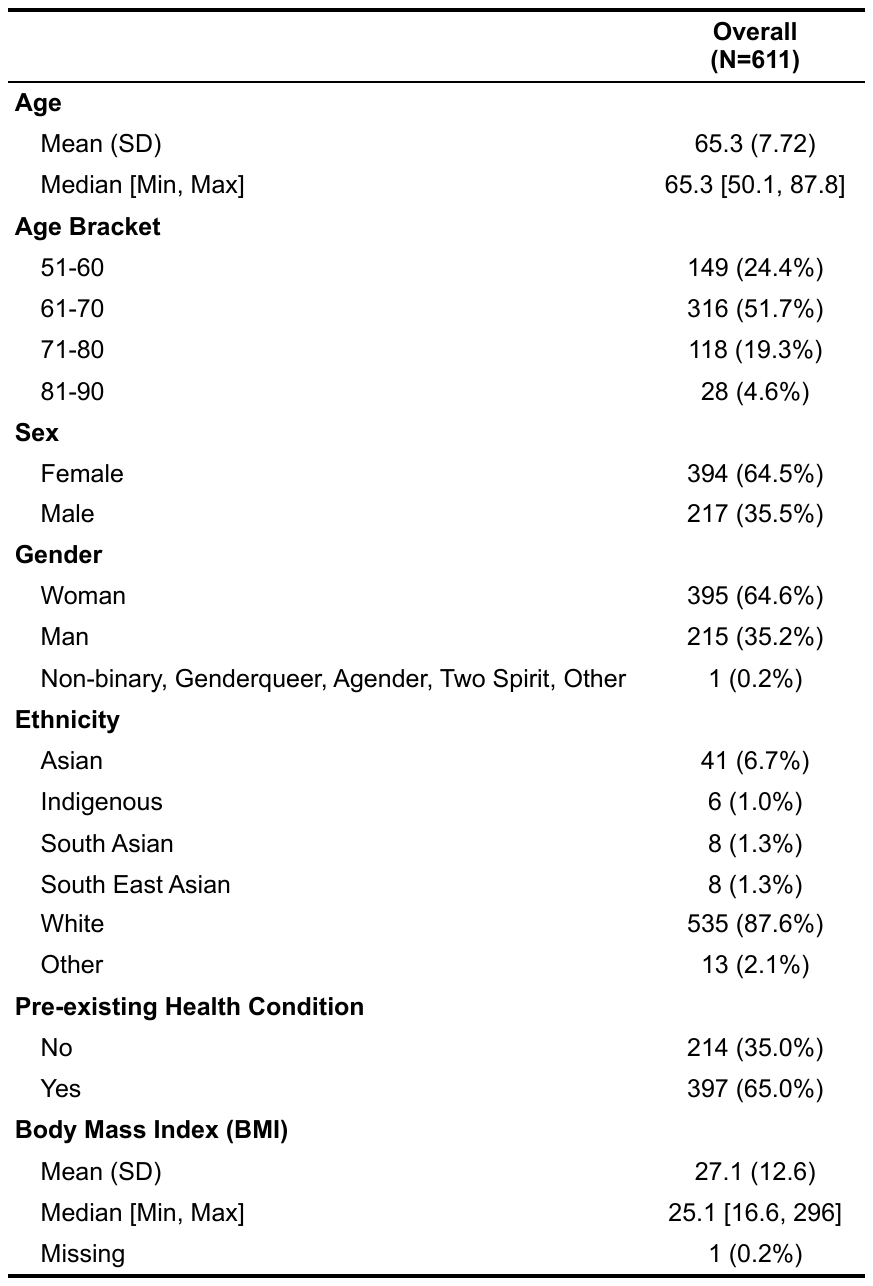

**Table S2. Vaccine type received by dosing regimen.**

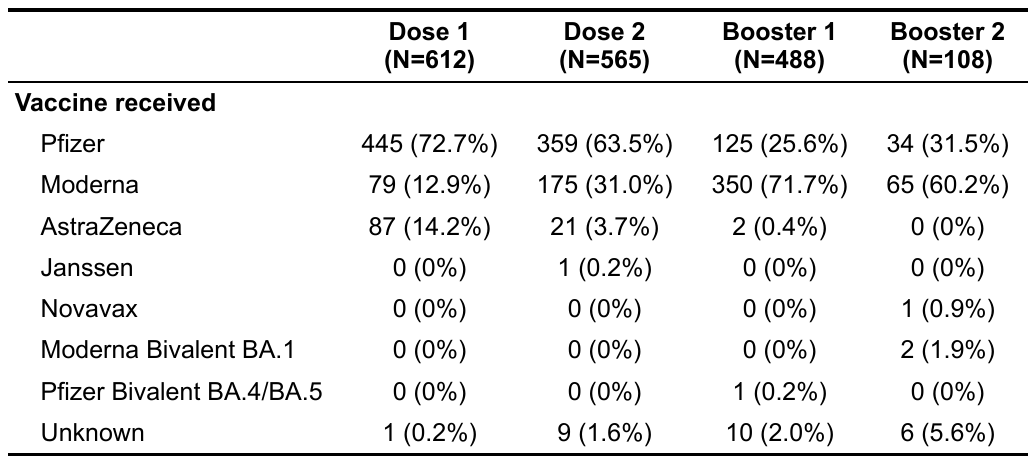

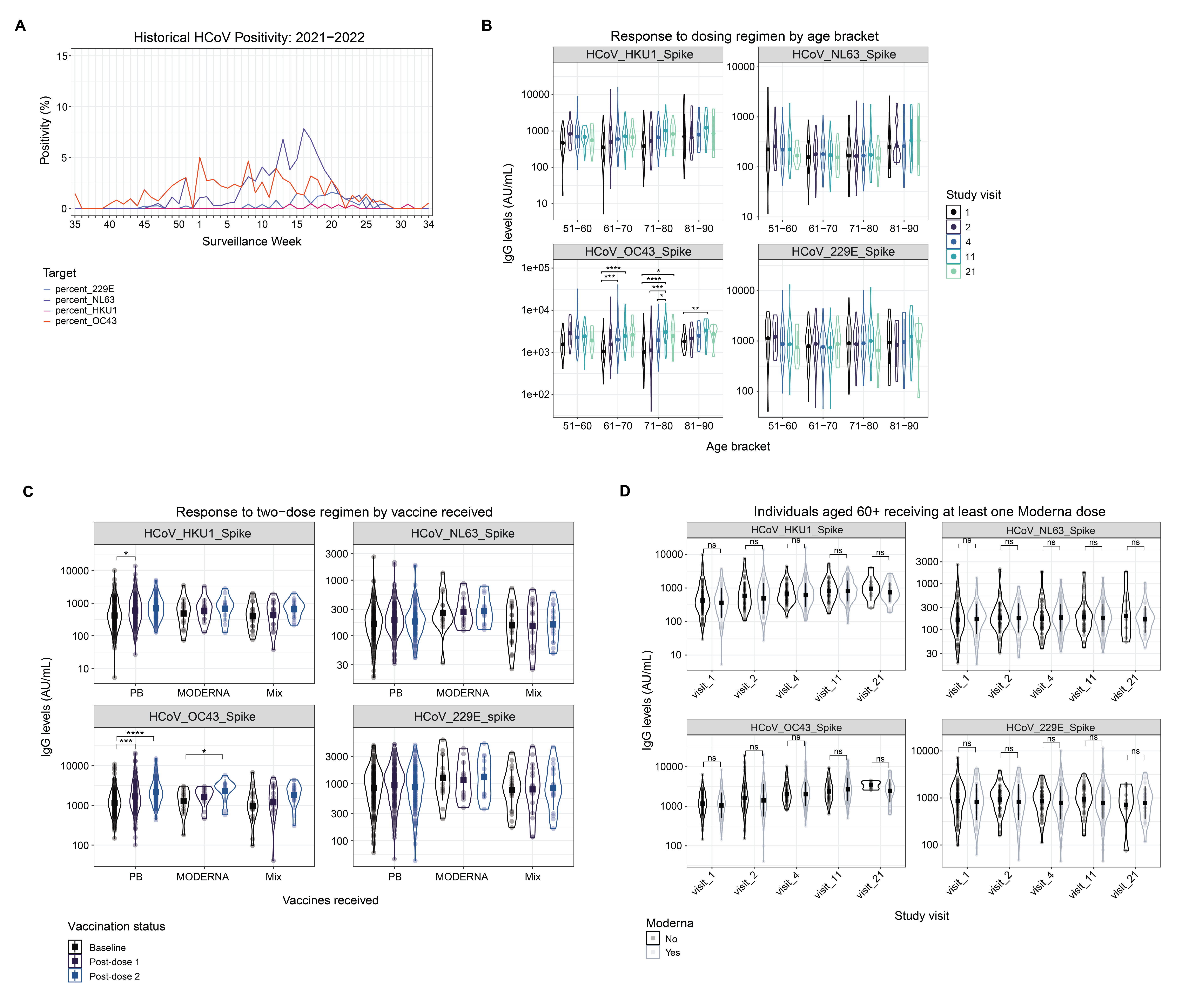

**Figure S2. Anti-HCoV antibody titres by vaccine status.** A) HCoV percent positivity rates by surveillance week during the September 2021 – August 2022 season. Colours represent individual HCoVs. B) Anti-endemic anti-S IgG at baseline and 1 month following each vaccine dose. Data presented by participant age bracket, coloured by study visit. C) Anti-endemic IgG titres in individuals aged 60+ at baseline, 1-month post-dose 1 and 1-month post-dose 2 by vaccine type received. Colours represent vaccination status. PB, BNT162b2. MODERNA, mRNA-1273. Mix, one of BNT162b2 and one of mRNA-1273 received in any order. D) Anti-endemic IgG titres in individuals aged 60+ at baseline and 1-month following each vaccine dose, comparing those who received at least one dose of mRNA-1273 throughout the study time course and those who did not.

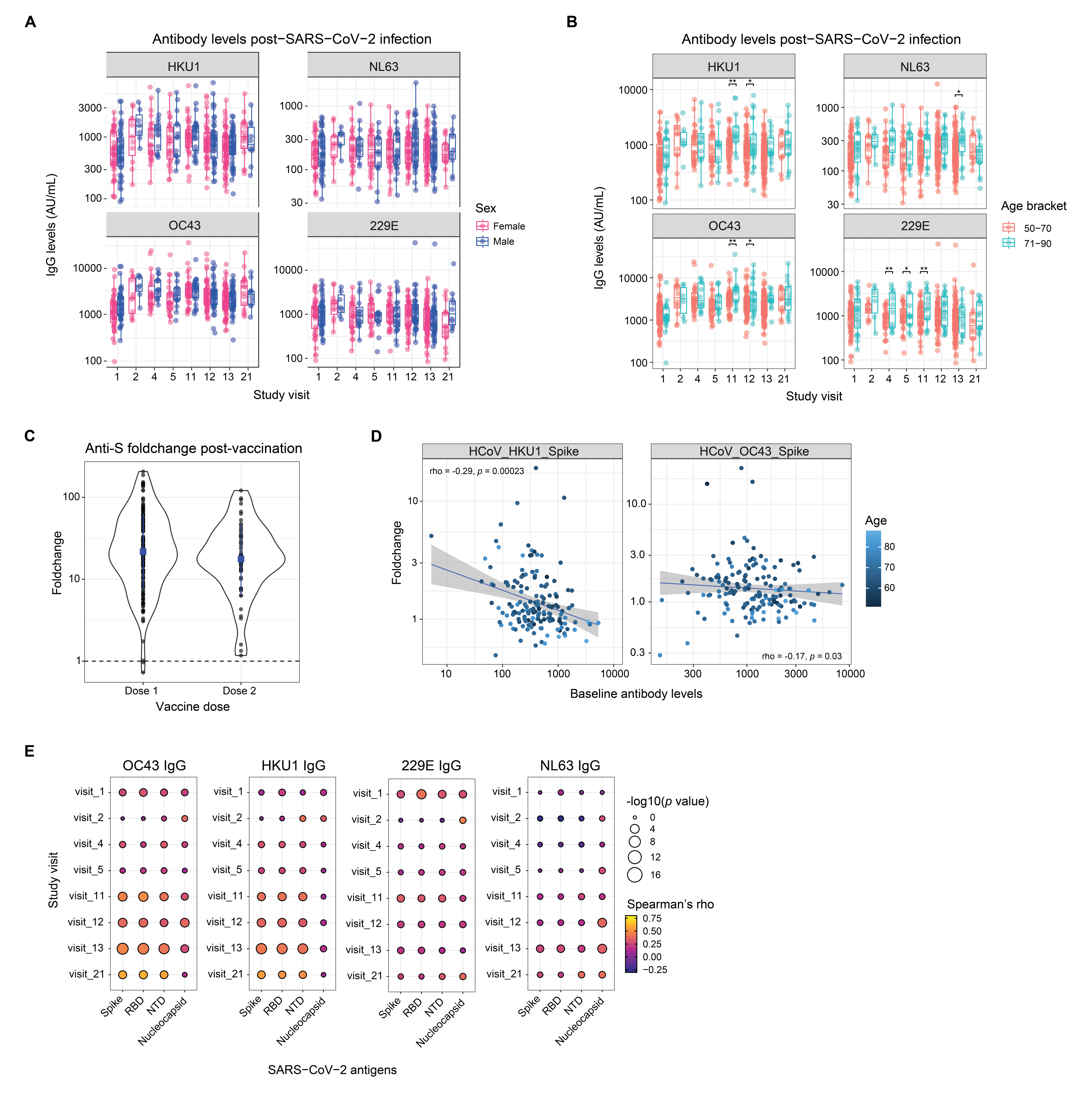

**Figure S3. Infection- and vaccine-induced immunity.** Anti-HCoV IgG levels in SARS-CoV-2 seropositive individuals at each study visit by participant A) sex and B) age bracket. C) Post-vaccination SARS-CoV-2 anti-S response at 1-month post-dose 1 and 1-month post-dose 2. D) Spearman correlation analysis of baseline anti-HKU1 and anti-OC43 antibody levels by their respective foldchanges post-vaccination. Colour represents participant age. Values represent Spearman’s rho. Line represents linear regression +/- 95% CI. E) Spearman correlation analysis of anti-HCoV antibody titres and antibodies specific to SARS-CoV-2 antigens by study visit in SARS-CoV-2 seropositive individuals. Size represents -log10(*p-*value). Colour represents Spearman’s rho.

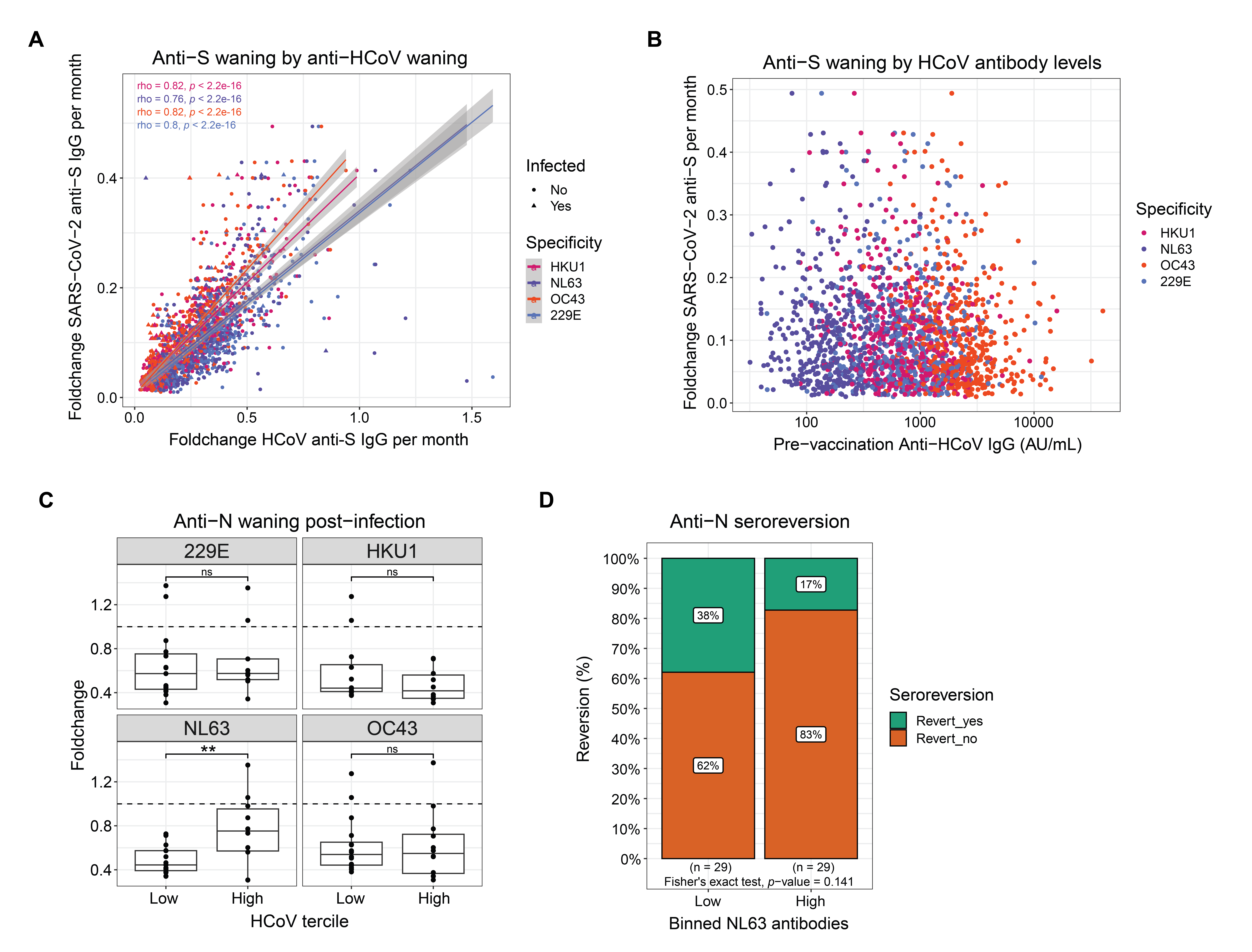

**Figure S4. Anti-S waning post-vaccination or infection.** A) Foldchange in SARS-CoV-2 anti-S antibody levels, normalized by time post-vaccination, plotted against the corresponding foldchange in anti-HCoV antibody levels. Statistics represent Spearman correlation analysis. Colour represents individual HCoVs. Symbol shape represents participant SARS-CoV-2 seropositivity. Values represent Spearman’s rho. Line represents linear regression +/- 95% CI. B) Foldchange in SARS-CoV-2 anti-S antibody levels, normalized by time post-vaccination, plotted against pre-vaccination levels of anti-HCoV antibodies. Colour represents individual HCoVs. C) Anti-N waning over a 3 month period post-first detected seropositivity. Waning is plotted as foldchange by HCoV antibody tercile at the time of infection. D) Relative proportion of individuals who demonstrated anti-N seroreversion from seropositive to seronegative anti-N status within the study time course following SARS-CoV-2 infection. Seroreversion is presented as a percentage of individuals by low and high NL63 antibodies at study start.

**Table S3. Multivariate analysis of SARS-CoV-2 infection risk throughout the study time course by baseline HCoV antibody tercile.**

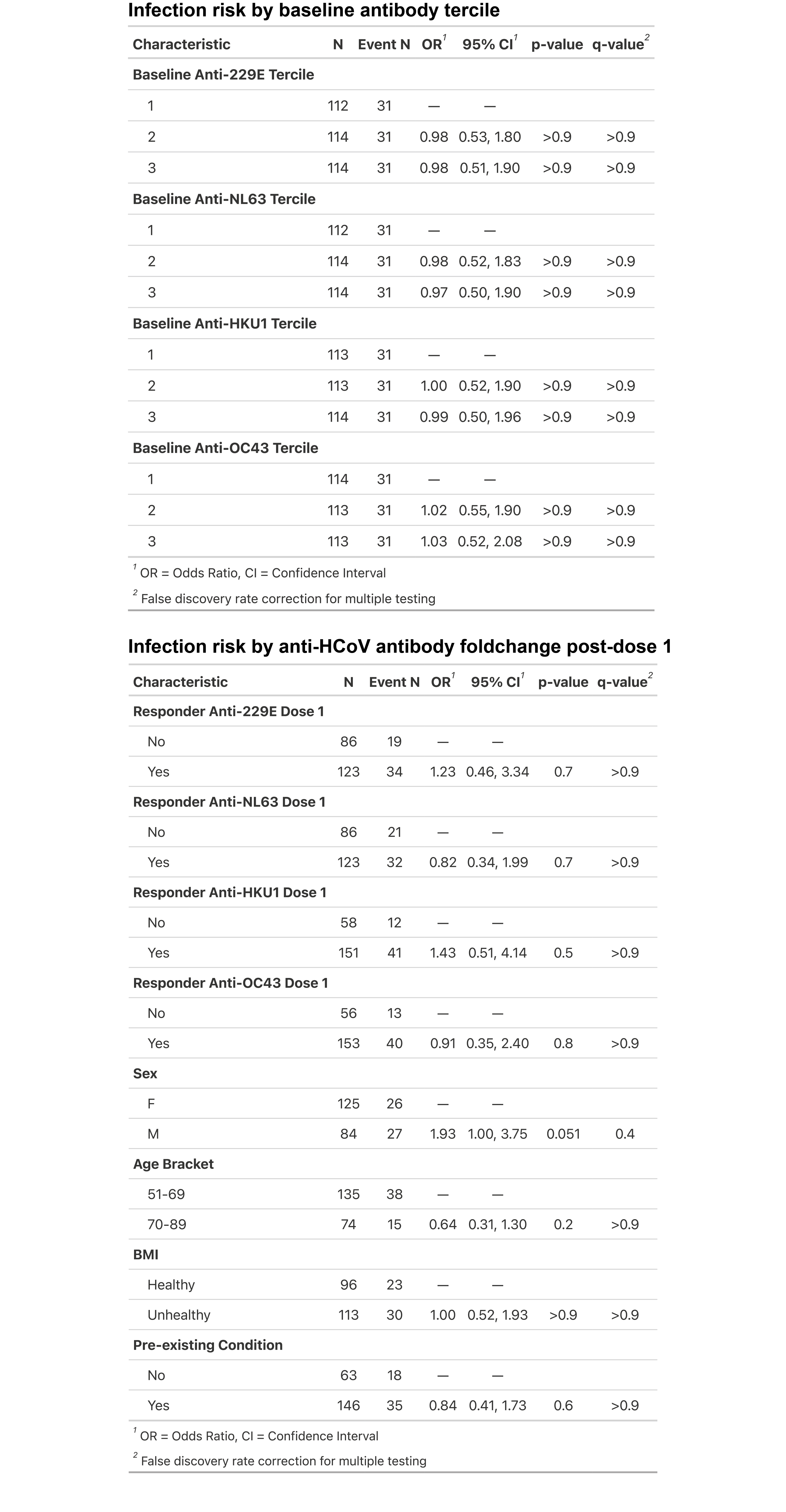

**Table S4. Multivariate analysis of SARS-CoV-2 infection risk throughout the study time course by post-dose 1 anti-HCoV antibody foldchange.**

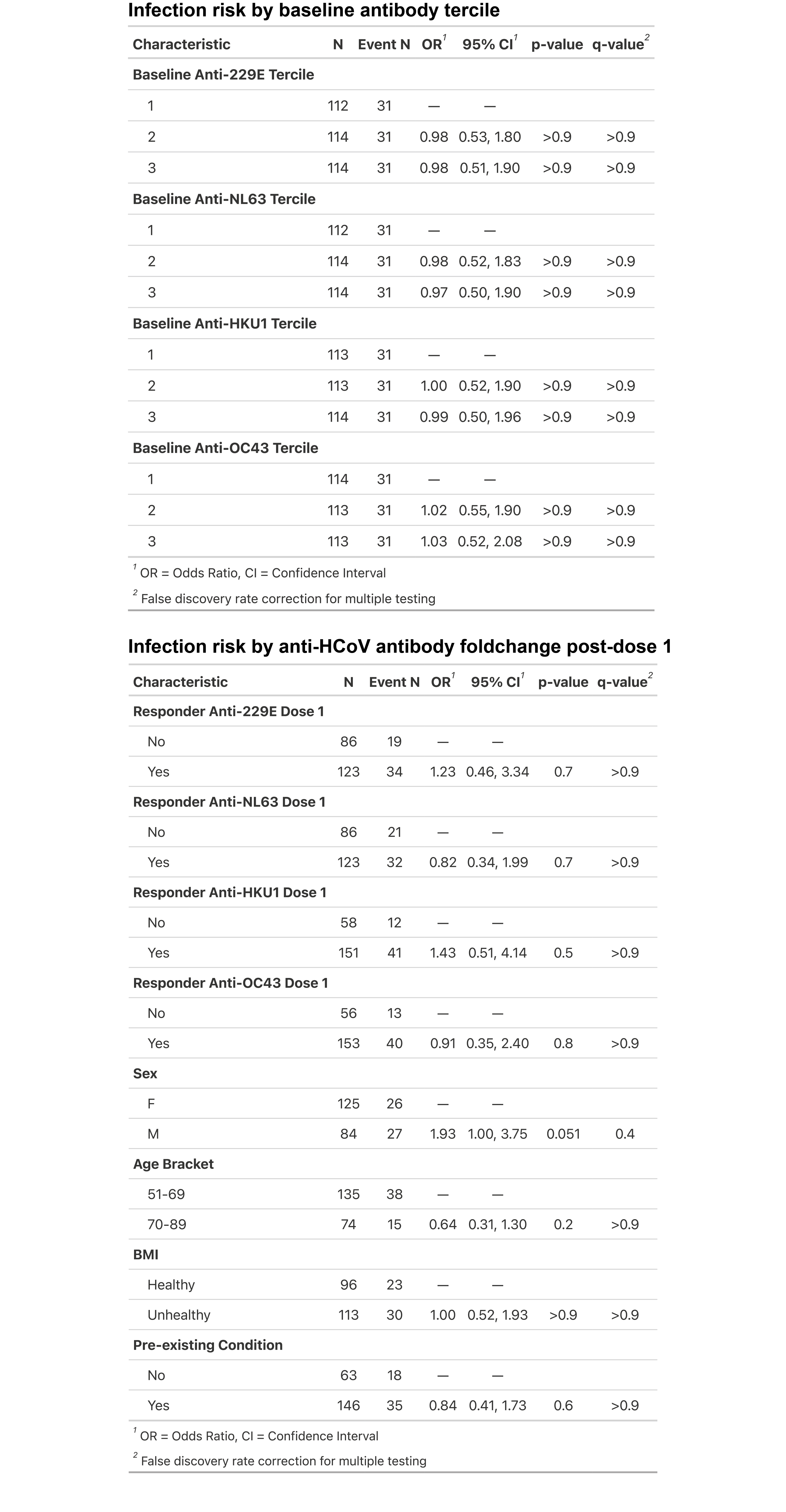

**Table S5. Multivariate analysis determining the odds of a high or low anti-HCoV antibody tercile by participant demographic factors.**

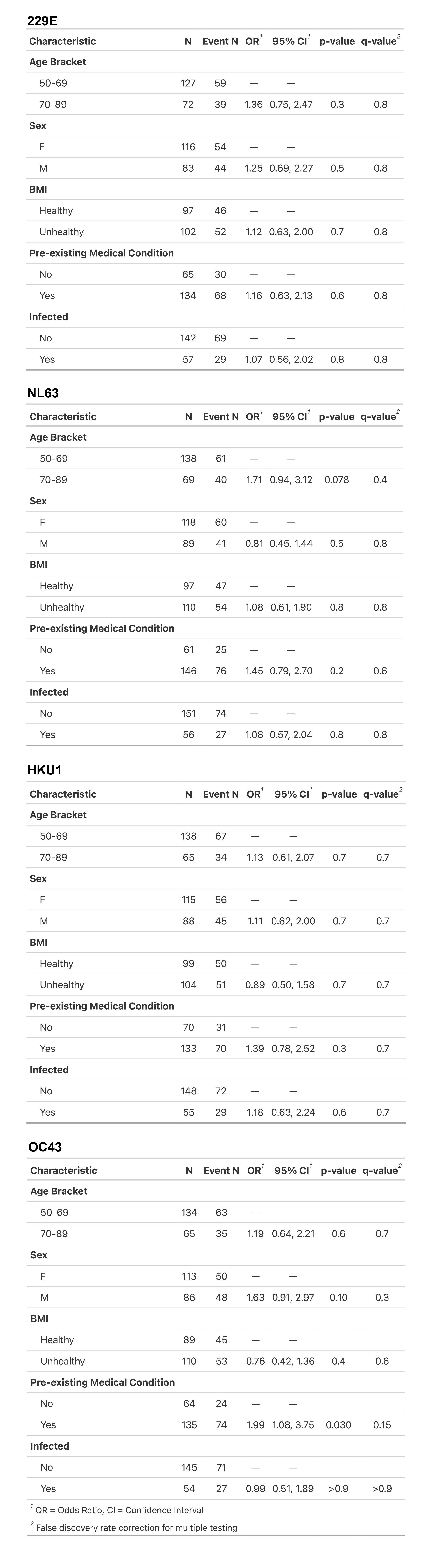

**Table S5 continued.**

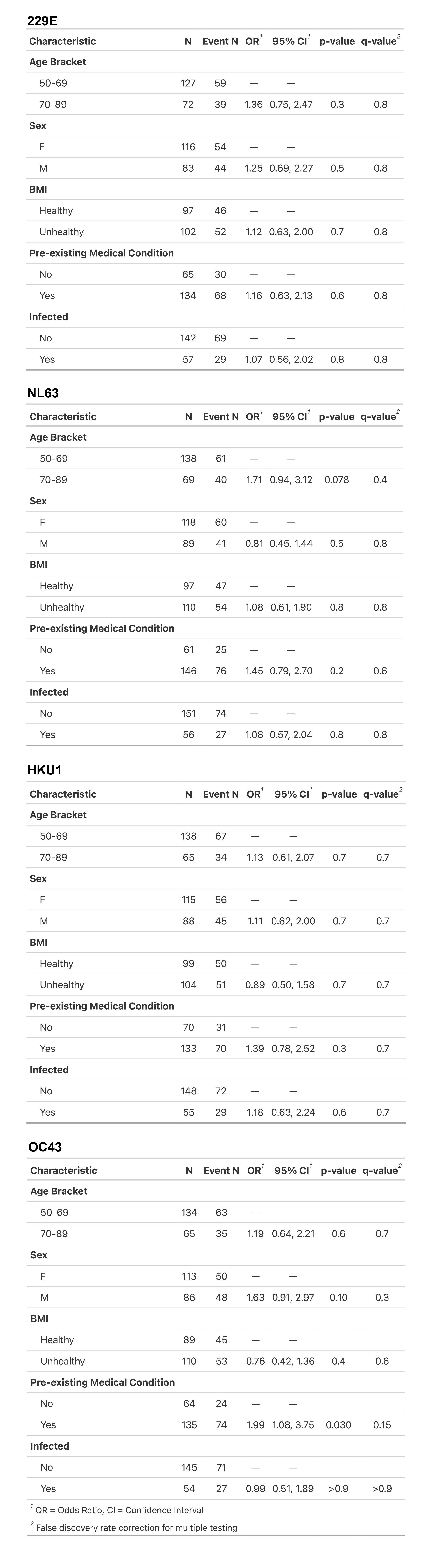

**Table S6. Correlation analysis of SARS-CoV-2 variant neutralization and anti-HCoV antibody levels.**

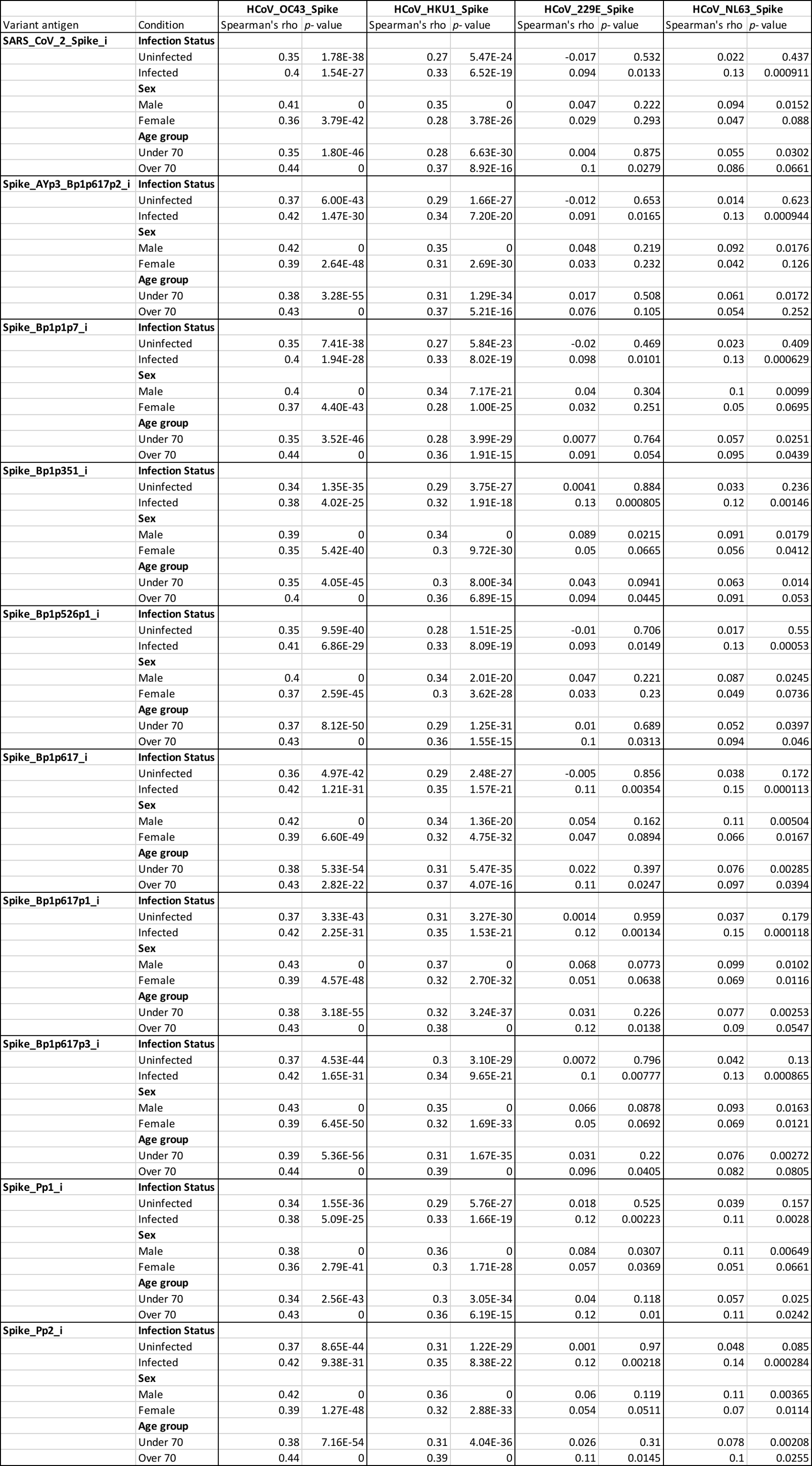

**Table S6 continued.**

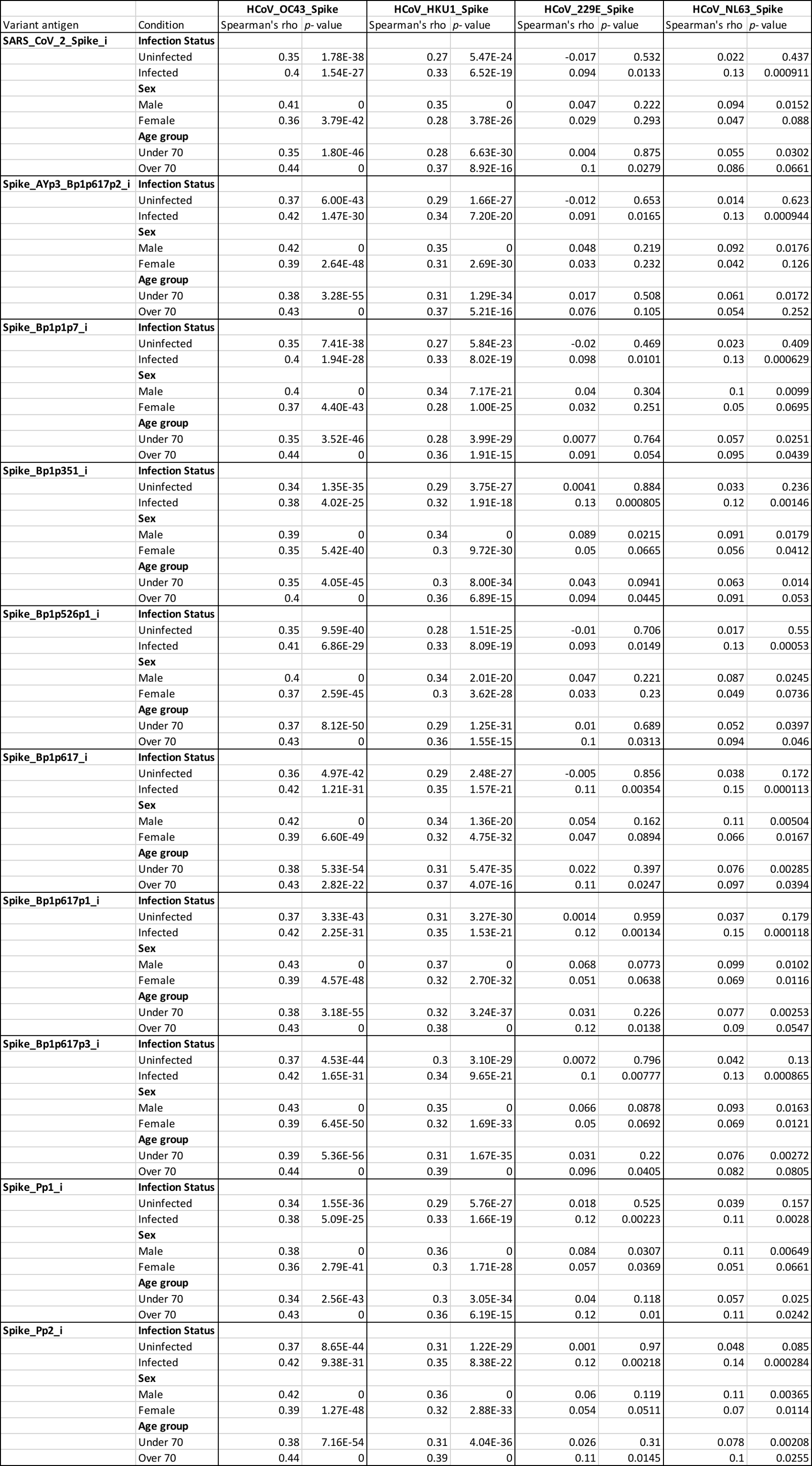
